# Identification of ovarian high-grade endometrioid-type tumors through multi-omics analysis: JGOG3025-TR2 study

**DOI:** 10.1101/2024.04.10.24305633

**Authors:** Shiro Takamatsu, R. Tyler Hillman, Kosuke Yoshihara, Tsukasa Baba, Muneaki Shimada, Hiroshi Yoshida, Hiroaki Kajiyama, Katsutoshi Oda, Masaki Mandai, Aikou Okamoto, Takayuki Enomoto, Noriomi Matsumura

**Author notes:** Corresponding author Noriomi Matsumura, Professor, Department of Obstetrics and Gynecology, Kindai University Faculty of Medicine, 377-2, Ohnohigashi, Osaka-Sayama, Osaka, 589-8511, Japan, (voice), (facsimile).

## Abstract

**Objective:** There is considerable interobserver variability in the differential diagnosis between ovarian high-grade endometrioid carcinoma (HGEC) and ovarian high-grade serous carcinoma (HGSC) due to their histopathological similarities. While the association between homologous recombination deficiency (HRD) and platinum sensitivity and PARP inhibitors is well established in HGSC, the molecular characteristics of HGEC remain unclear.

**Methods:** Fresh-frozen tissue samples from 15 ovarian HGECs and 274 ovarian HGSCs morphologically diagnosed by central pathology review in the Japanese Gynecological Oncology Group (JGOG) were subjected to targeted DNA sequencing, RNA sequencing, DNA methylation array, and SNP array analysis. Tumors were classified by unsupervised clustering based on copy number variation signatures. External datasets including 555 ovarian HGSCs and 287 endometrial high-grade carcinomas from The Cancer Genome Atlas project (TCGA-OV and TCGA-UCEC) were also analyzed.

**Results:** Four distinct groups were identified in the JGOG cohort. C1 (n=41) showed *CCNE1* amplification and poor survival. C2 (n=160) and C3 (n=59) showed a high frequency of *BRCA* alterations with moderate and low aneuploidy, respectively. C4 (n=22) was characterized by favorable survival, higher frequency of HGEC, absence of both *BRCA* alteration and *CCNE1* amplification, and low levels of HRD score, ploidy, intra-tumoral heterogeneity, cell proliferation rate, and *WT1* gene expression. Additionally, C4 exhibited a normal endometrium-like DNA methylation profile and was defined as an “HGEC-type” tumor. The HGEC-type tumors were also identified in TCGA-OV and TCGA-UCEC.

**Conclusions:** Ovarian HGEC-type tumors exhibit non-HRD status, a favorable prognosis, and endometrial differentiation, and may comprise a subset of tumors diagnosed as HGSC.

## Introduction

Ovarian cancer has the highest mortality rate among gynecologic malignancies and its incidence is increasing in many countries around the world (#1, #2). Epithelial ovarian cancer, which accounts for approximately 90% of ovarian malignancies, includes several major histologic types, including high-grade serous carcinoma (HGSC), low-grade serous carcinoma, endometrioid carcinoma, clear cell carcinoma, and mucinous carcinoma (#2). These different histologic types are related to molecular characteristics, including gene expression, DNA methylation, gene mutations, and copy number variations, which influence clinical presentation and response to drug treatment (#4, #5, #6, #7, #8, #9). HGSC is the predominant histologic type, accounting for approximately 70% of epithelial ovarian cancers, and most cases are diagnosed as advanced cases with peritoneal dissemination (#2). On the other hand, endometrioid carcinoma, which is the most common histologic type of endometrial cancer, represents a minor histologic type, accounting for only about 10% of epithelial ovarian cancers (#2, #10, #11).

Similar to endometrial cancer, ovarian endometrioid carcinomas are classified into low-grade (LGEC; grade 1 or 2) and high-grade (HGEC; grade 3) based on the proportion of solid components, with the latter having a poorer prognosis than the former (#12). Earlier studies reported that HGEC were indistinguishable from HGSC by gene expression profile (#13, #14) and had *TP53* mutation, which is commonly observed in HGSC, but lacked gene mutations such as *KRAS, PIK3CA, PTEN*, and *CTNNB1*, which are occasionally observed in LGEC. However, subsequent studies showed that some of the morphologic features previously thought to be characteristic of HGEC, such as SET (Solid, pseudo-Endometrioid, and Transitional cell carcinoma-like morphology) features, could be considered as HGSC (#15, #16). As a result, a significant proportion of tumors previously diagnosed as HGEC are now diagnosed as HGSC (#17), and HGEC is recognized as an uncommon tumor.

In the past decade, comprehensive molecular studies, including The Cancer Genome Atlas (TCGA) or the International Cancer Genome Consortium (ICGC) projects, have shown that more than half of HGSCs have homologous recombination deficiency (HRD) and that gene mutations in *BRCA1/2* and characteristic chromosomal changes of HRD are biomarkers of sensitivity to platinum and PARP inhibitors (#9). Previously, clinical trials testing the efficacy of PARP inhibitors in ovarian cancer included only HGSC cases (#18, #19), but more recently, HGEC cases have also been included (#20, #21, #22, #23). This is because the molecular features specific to HGECs that distinguish HGSCs from HGECs have not been fully elucidated.

In the present study, we analyze the multi-omics dataset including both HGEC and HGSC, identify the molecular characteristics of HGEC, and demonstrate the inclusion of HGEC-type tumors among tumors diagnosed as HGSC. The results will advance the molecular classification of ovarian cancer and personalized medicine.

## Material and Methods

### Analysis of the JGOG3025 and JGOG3025-TR2 (JGOG-TR2) datasets

#### JGOG-whole cohort

In the JGOG3025 study, the Japanese Gynecologic Oncology Group (JGOG) collected clinical information and targeted DNA sequencing data for 51 genes in 710 cases of epithelial ovarian cancer, including 298 high-grade serous carcinoma (HGSC) and 24 high-grade (grade 3) endometrioid carcinoma (HGEC) (#7).

#### Central pathological review

As previously reported (#7), the central pathological review (CPR) was performed by three independent pathologists (Professor Yuko Sasajima, Professor Miki Kushima, and Dr. Masaharu Fukunaga) assigned by the JGOG. Evaluation and diagnosis were based on the WHO classification of tumors of female reproductive organs using only the representative hematoxylin and eosin staining (HE) slides of tumors without any reference to the results of immunohistochemistry (IHC) staining.

#### Microsatellite instability (MSI) high and tumor mutational burden (TMB) high tumor

MSI status was estimated from raw targeted DNA sequencing data using MSIsensor2 (https://github.com/niu-lab/msisensor2). Genomic regions of the panel contained 18 microsatellite sites. According to the previously reported threshold of 30% (#24), samples with microsatellite instability at 6 (33%) or more sites were defined as MSI-high (Supplementary figure S%1%AB). TMB-high tumors were defined as tumors that had equivalent number of mutations as MSI-high tumors (Supplementary figureS%1%C).

#### JGOG-TR2 cohort

Fresh-frozen tumor tissues from 274 and 15 cases diagnosed as stage II or higher HGSC and HGEC in the CPR were submitted to SNP array (Affymetrics OncoScan), total RNA-sequencing, and DNA methylation array (Illumina EPIC) analyses. Sample preparation and pretreatment for these analyses were described in detail previously (#25).

#### Gene mutation profile and LOH status of gene mutations in HRR-related genes

For all detected variants, the following database information was added using ANNOVAR (#26): Clinvar (2023Jan05), InterVar (2018Mar25), dbNSFP, dbscsNV, dbSNP, gnomAD, 1KGP Genomes Project (1000g2015aug), ExAC, ESP6500, and 54K Japanese custom reference from jMorp (8.3KJPN). In addition, the latest OncoKB (#27) and BRCA Exchange (#28) annotations as of November 2023 have been added. Variants with VAF < 5% and those with population allele frequency > 1% were excluded. Variants annotated “benign” in Clinvar, InterVar, oncoKB, or BRCAexchange were excluded, while those “pathogenic” or “oncogenic” were retained. Missense mutations were retained if they were considered loss-of-function type in multiple functional prediction tools using dbSNFP annotations. Splice site mutations were included if they were considered loss-of-function type in the dbscSNV annotations.

The following were defined as genes associated with homologous recombination repair pathways: *ATM, ATR, ATRX, BAP1, BARD1, BLM, BRCA1, BRCA2, BRIP1, CHEK1, CHEK2, FANCA, FANCC, FANCL, MLE11, NBN, PALB2, RAD 21, RAD50, RAD51, RAD51B, RAD51C, RAD51D, RAD52, RAD54L, RECQL4*, and *XRCC2*. Since it was unclear whether mutations in these genes cause HRD in the absence of locus-specific loss-of-heterozygosity (LOH) (#7), variants were retained as significant only when the copy number of the minor allele at the locus was 0.

#### Copy number variations

As previously reported (#25), segmented allele-specific copy number variations, estimated tumor ploidy, and purity were calculated using ASCAT (#29) from the SNP array data. For samples whose tumor purity was not determined to be in the range of 0.20 to 0.95 in the initial analysis, optimal purity and ploidy were adjusted manually with reference to the generated “sunrise plots”. Three samples for which the optimal values could not be determined from the plots were excluded from the analysis.

From the ASCAT output, the 48-channel CNV signatures were obtained using SigProfilerMatrixGenerator (#30) and the HRD score was calculated as the sum of the LST, the TAI, and the LOH scores using scarHRD (#31). *CCNE1* amplification was defined when the total copy number of the *CCNE1* gene region was greater than or equal to 6.

#### Hierarchical clustering and determination of the number of clusters

After the 48 CNV signature values per sample were normalized by dividing by the total number of segments per sample, Ward’s hierarchical clustering was performed. We determined the number of clusters to be four based on the average silhouette scores for different numbers of clusters (Supplementary figure S%2%AB), the distribution of samples in PCA (Supplementary figure S%2%C) and UMAP (Supplementary figure S%2%D), and other molecular differences among the assigned clusters.

#### BRCA1 methylation silencing

As previously reported (#25), *BRCA1* promoter CpG island probes were selected when the mean beta was <0.3 and the Spearman correlation between mRNA expression and beta value was rS <−0.1, P value <0.01. Samples were annotated as methylation silenced when all the selected probes met the criteria of beta value >0.3 and mRNA expression <30%.

#### Determination of HRD score cutoff

As previously reported (#25), using the ROC curve and Youden’s index, the optimal cutoff of the HRD score that best discriminates samples with *BRCA1/2* mutations with LOH and *BRCA1* methylation was determined as 67 in the JGOG-TR2 dataset.

#### Differentially gene expression analysis between the C4 vs the C1-C3

We compared gene expression profiles between the C4 and the C1-C3 tumors using limma+voom (#32, #33) and DESeq2 (#34). After filtering low expressed genes (Supplementary figure %3%), genes with log2 fold change >1(UP) or <-1 (DN) and adjusted P-value<0.01 were defined as differentially expressed genes (Supplementary figure %3%). 147 upregulated genes and 161 downregulated genes that were commonly identified by the two methods were determined as the JGOG-C4 gene signature (Supplementary table 1). We calculated the JGOG-C4 expression score by subtracting enrichment score of the down-regulated genes from that of the upregulated genes using ssGSEA (#35).

### EM/FT methylation score and EM/FT silencing score

To evaluate differences in genome-wide methylation status associated with cell differentiation between normal endometrium (EM) and fallopian tubes (FT), we developed the EM/FT methylation score. Using whole genome bisulfite sequencing data in a previous study (GSE1864888) (#36), we analyzed the differentially methylated regions (DMRs) between 3 normal endometrial epithelium (EM) and 3 normal fallopian tube epithelium (FT) using “find markers” function in wgbs_tools (#36). 6626 genomic regions were identified as hyper-methylated regions in EM than in FT (EM>FT regions) and 5372 genomic regions were identified as FT>EM regions. Subsequently, using Methyl-ssRSEA (#37), which accepts beta values from the Infinium MethylationEPIC (EPIC) or Infinium HumanMethylation450 BeadChip (HM450) as input, the EM/FT methylation score was calculated as the subtraction of FT>EM region enrichment score from EM>FT region enrichment score.

To assess the gene silencing related to the EM/FT methylation, we developed the EM/FT silencing score. Using annotator (#38), 432 promotor CpG islands of 208 genes (EM>FT silencing genes, Supplementary table 2) were identified in the EM>FT regions and 236 promotor CpG islands of 106 genes (FT>EM silencing genes, Supplementary table 2) in the FT>EM regions. The EM/FT silencing score was defined as the subtraction of enrichment scores of these genes (EM/FT – FT/ET) using ssGSEA (#35).

To examine the EM/FT methylation score, we obtained publicly available datasets of genome-wide DNA methylation array, including GSE51820 (#5), GSE72021 (#42), GSE155760 (#43), GSE168930 (#44), GSE226823 (#45) (Supplementary table 3). Similarly, to test the EM/FT silencing score, we obtained previously studied gene expression profiles from GSE2109, GSE6008 (#46), GSE19539 (#47), GSE44104 (#48), GSE65986 (#49), and the ICON7 study (#50) (Supplementary table 3).

### TCGA-OV/UCEC data analysis

We obtained multi-omics datasets of The Cancer Genome Atlas (TCGA) project for high-grade serous ovarian cancer (TCGA-OV) (#39) and uterine corpus endometrial cancer (TCGA-UCEC) (#40) from the GDC portal (#41) (Supplementary table 3). From segmented allele-specific copy number variations processed by ASCAT (#29), 48-channel CNV signatures and HRD scores were calculated using SigProfilerMatrixGenerator (#30) and scarHRD (#31) in the same way as the JGOG-TR2. Somatic mutation profiles were downloaded from the GDC portal and mutations with “Oncogenic” annotation in the latest OncoKB (#27) were retained. Processed gene expression TPM values and DNA methylation data were downloaded from the GDC portal (#41). Using gene expression and gene promotor-based beta values in *BRCA1* and *MLH1*, samples whose beta-value > 0.3 and expression < 30% was annotated as methylation silenced. Mass spectrometry-based protein expression for TCGA-OV samples studied in the Clinical Proteomic Tumor Analysis Consortium (#51) was obtained from cBioPortal (#52).

In TCGA-UCEC, cases with histopathologic diagnoses of serous carcinoma and grade 3 endometrioid carcinoma were selected. Cases with mutations in *POLE* and four MMR-related genes, including *MLH1, MSH2, MSH6, PMS2*, and with *MLH1* methylation were defined the POLE/dMMR subtypes.

Most of the TCGA-OV samples were analyzed using the Infinium HumanMethylation27 (HM27), while most of the TCGA-UCEC samples using HM450K. Since HM27K probes are designed to mainly cover known gene promoter CpG islands and do not cover the whole genomic regions as comprehensively as HM450 (#53), the EM/FT methylation score using Methyl-ssRSEA (#37) was not calculated for TCGA-OV samples.

### Machine learning subtype prediction based on CNV signatures

48-channel CNV signature values from TCGA-OV were adjusted to those from the JGOG-TR2 cohort using Combat_seq (#54). In TCGA-UCEC, because the SNP array platform was identical to TCGA-OV, CNV signature values of these two datasets were simply combined and then adjusted to those from the JGOG-TR2 cohort using Combat_seq.

Using adjusted CNV signatures as features and the C1-C4 subtypes as labels, five different classifier models, including k-nearest neighbor, support vector machine, random forest, multilayer perceptron, XGBoost, were trained in the JGOG-TR2 cohort. All the classifiers were used in default parameters of scikit-learn packages, except for the random forest classifier, where the maximum depth of the tree was set as 3 to prevent over-learning.

We set stringent criteria in the machine learning approach, where results were only adopted if they were consistent across four or more of five classification methods.

### Analysis in the Surveillance, Epidemiology, and End Results (SEER) database

From the database of 22-registry SEER Research limited field data (November 2022 submissions), we selected cases with site recode (ICD-O-3 2023 Revision) ‘Ovary’ and histological diagnosis (ICD-O-3 Hist/behav) ‘8380/3: Endometrioid carcinoma’ and examined the number of cases stratified by year of diagnosis and histological grade (‘Grade Recode (thru 2017)’ or ‘Grade Pathological (2018+)’).

### Statistical analysis

All statistical analyses in this study were performed using Python (3.8.8). For comparison between two groups, T-test was used for continuous values and Chi-square test for binary values using SciPy (1.7.2). Machine-learning analyses were performed using Scikit-learn (1.0.1) and XGBoost (v). Survival analyses including the Kaplan–Meier curve, the multivariate log-rank test, and the Cox proportional hazard regression analysis were performed using Lifelines (0.26.3). A P value <0.05 was considered statistically significant.

### Data and codes availability

Processed data and analysis codes to reproduce the results in this study are available in the GitHub project page (https://github.com/shirotak/JGOG_HGEOC). Genomic data of the JGOG3025-TR2 cohort, including RNA-seq, SNP array, and DNA methylation array are available from SRA (PRJNA1092599) and NCBI-GEO (GSE263455***). Targeted sequencing data and clinical information are available upon reasonable request to JGOG (, https://jgog.gr.jp/en/index.html).

Controlled access data for the TCGA cases were obtained through dbGaP (access permission phs000178). Gene expression data for the ICON7 cases were obtained through the European Genome-Phenome Archive (accession number EGAS00001003487).

*** The dataset will be made available after publication of the paper

## Results

### Molecular profiles based on pathological diagnosis

In the JGOG-whole cohort, ovarian HGEC was diagnosed in 16.9% (24/142) of cases with ovarian endometrioid carcinoma. The percentage was consistent with the ratio of HGEC diagnosis calculated from the latest SEER database (Supplementary figure S%4%).

Clinical characteristics of the JGOG-TR2 study cohort are shown in Table1. Targeted DNA sequencing, RNA sequencing, DNA methylation array, and SNP array analyses were performed on 289 fresh-frozen tumor samples with pathological diagnosis of HGEC (n=15) and HGSC (n=274) (see Methods).

In targeted DNA sequencing, gene mutations commonly observed in ovarian endometrioid carcinoma (#55, #56), including *ARID1A, PTEN, PIK3CA*, and *KRAS*, were detected in HGEC (Supplementary figure S%5%A). The Homologous Recombination Deficiency (HRD) score was higher in HGSC than HGEC (Supplementary figure S%5%B, P=.028). Estimated ploidy was higher in HGSC than HGEC (Supplementary figure S%5%C, P=1.6 × 10^−4^). The MATH score, a measure of intratumor heterogeneity calculated from the mutant allele frequency profile, was higher in HGSC than HGEC (Supplementary figure S%5%D, P=5.5 × 10^−5^). In the analysis of the TCGA-OV data, *WT1* gene expression was significantly positively correlated with WT1 protein expression, a commonly used diagnostic marker for serous subtype (Spearman r=.65, P=1.3x10^−16^, Supplementary figure S%6%). Therefore, we examined *WT1* gene expression and found that it was higher in HGSC than HGEC (Figure S%5%E, P=3.1 × 10^−4^).

In the centralized pathological review (CPR), the pathological diagnosis was solely based on morphological findings on hematoxylin-eosin (HE) stained slides, without any reference to the results of immunohistochemical (IHC) staining, which was performed at the initial treatment institutions when necessary. The concordance of histologic diagnosis of HGEC between CPR and initial treatment institution (ITI) was low; only six tumors were common for HGEC diagnosis between CPR and ITI (Figure %5%F).

The significant discrepancy observed in the pathologic diagnosis of HGEC between CPR and ITI was consistent with previous reports that it is difficult to differentiate HGEC from HGSC based on histopathological findings (#17, #57). Therefore, we aimed to classify the JGOG-TR2 tumors based on molecular features rather than morphological findings. Since chromosomal instability is one of the most characteristic molecular features of HGSC (#58), we focused on CNV signature-based analysis (#59).

### Identification of four tumor subtypes based on CNV signatures

Unsupervised hierarchical clustering based on 48-channel CNV signatures (see Methods) identified four different clusters with distinctive genomic features (Figure 1A, Supplementary figure S%7%A). The C1 cluster (n=41) showed a high *CCNE1* amplification rate (P=8.1x10^−16^), low frequency of *BRCA1/2* alterations (*BRCA1/2* mutation with locus-specific LOH and *BRCA1* methylation silencing) (P=3.2x10^−6^), complete absence of HGEC, moderate HRD score (C2/C3 > C1: P=2.1x10^−5^, C1> C4: P=1.5x10^−41^), and high ploidy (P=2.7x10^−53^), which is known to be associated with *CCNE1* amplification (#60). Both the C2 (n=160) and the C3 (n=59) clusters showed a high *BRCA1/2* alteration rate (P=4.9x10^−10^) with a high HRD score (P=7.1x10^−22^), but the C3 showed very low ploidy (C2>C3: P=1.4x10^−47^). The C4 cluster (n=22) showed distinctive features including significantly higher frequency of cases diagnosed with HGEC (P=1.8x10^−6^), lower *TP53* mutation rate (P=3.0x10^−8^), complete absence of *BRCA* alterations (P=4.0x10^−4^) or *CCNE1* amplification (P=.033), higher frequency of *PIK3CA/ARID1A/KRAS/PTEN* mutations (P=4.2x10^−7^), lower HRD score (P=4.5x10^−42^), lower ploidy (P=6.4x10^−5^), lower MATH score (P=.0011), and lower *WT1* gene expression (P=8.1x10^−10^) than the other groups.

**Figure 1).**
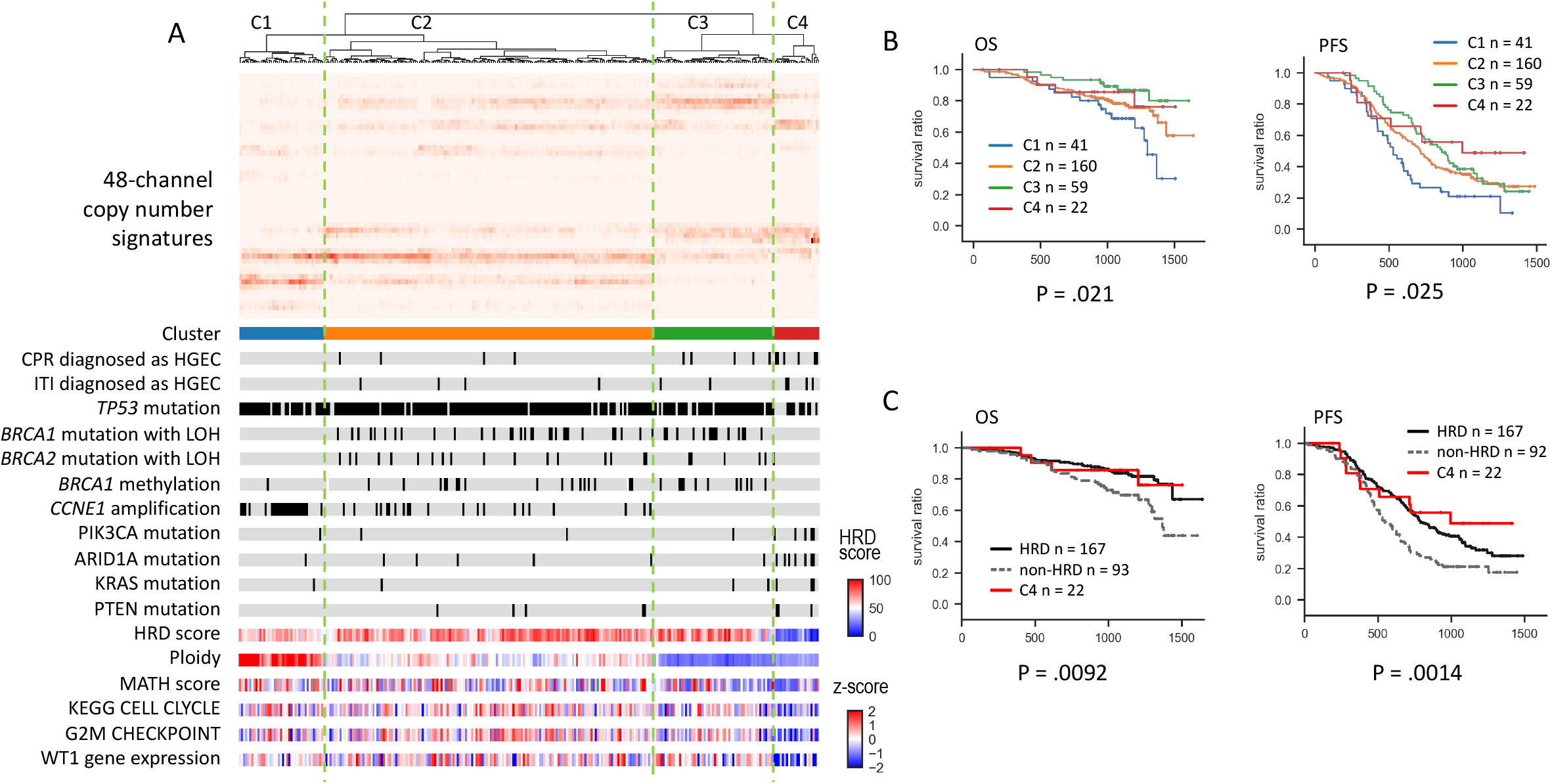
Four tumor subtypes based on CNV signatures in the JGOG3025-TR2 cohort. A) Molecular profiles. Unsupervised clustering based on 48-channel CNV signatures revealed the four tumor subtypes (Supplementary figure 2). The C1 showed a high *CCNE1* amplification rate, a low frequency of BRCA1/2 alterations, a complete absence of HGEC, moderate HRD score, and high ploidy. Both C2 and C3 showed high BRCA alteration rates and high HRD scores, while C3 exhibited very low ploidy. C4 showed a low *TP53* mutation rate, a complete absence of BRCA alterations and *CCNE1* amplifications, and an extremely low HRD score. B) Comparison of overall survival (OS) (left) and progression-free survival (PFS) (right) among the four subtypes. C) Comparison of OS (left) and PFS (right) between HRD, C4, and non-HRD without C4. While all the C4 cases were classified into non-HRD, they tended to have as favorable outcomes as HRD cases.

KEGG_CELL_CYCLE and G2M_CHECKPOINT gene expression scores calculated using ssGSEA were significantly lower in the C4 cluster than the others (P=1.9x10^−7^ and P=1.1x10^−9^), suggesting a lower cell proliferation rate of the C4 tumors. Collectively, the C4 tumor subtype was considered to be a molecularly determined “HGEC-type” tumor.

When compared to tumors pathologically diagnosed as HGEC in CPR (n=15), ITI (n=16), and either of them (n=25), the C4/HGEC-type tumors tend to have more distinctive molecular features from the other tumors (Supplementary figure S%7%B).

Overall survival (OS) and progression-free survival (PFS) were significantly different among the four subtypes (P=.021 and P=.025, Figure1B), even when analyzing within pathologically diagnosed HGSC samples (Supplementary figure S%7%C). The C1 showed the worst prognosis and was considered a previously known *CCNE1* amplification subtype in HGSC (#61). When tumors were divided into HRD and non-HRD types according to *BRCA1/2* alterations and the HRD score (see Methods), while all the C4 cases were classified into non-HRD, they tended to have a better prognosis than other non-HRD cases (Figure 1C). The results were similar when the analysis was limited to pathologically diagnosed HGSC tumors (Supplementary figure S%7%D).

### Genome-wide DNA methylation analysis associated with cell differentiation of endometrium and fallopian tubes

To investigate genome-wide DNA methylation associated with cell differentiation between uterine endometrium (EM) and fallopian tubes (FT), we utilized whole genome bisulfite sequencing data from a previous study (#36) to analyze differentially methylated regions (DMRs) between EM and FT, and developed the EM/FT methylation score (Supplementary figure %8%A).

In a DNA methylation array dataset for normal EM (n=13) and FT (n=11) samples (GSE155760) (#43), the EM/FT methylation score was significantly higher in EM than in FT (P= 7.2x10^−5^, Figure 2A). In the JGOG-TR2, the EM/FT methylation score was significantly higher in CPR-diagnosed HGECs (P=1.4x10^−4^) than in HGSCs, but the difference was more striking between the C4 tumors and the others (P=2.7x10^−18^) (Figure 2B). In another DNA methylation array dataset for ovarian cancer (GSE226823) (#45), including 60 HGSC, 19 endometrioid carcinoma, and 48 clear cell carcinoma samples, the EM/FT methylation score was significantly higher in endometrioid carcinoma than in HGSC (Figure 2C, P=2.1x10^−17^). This difference was consistently observed in other genome-wide DNA methylation array datasets (Supplementary figure %8%B). Interestingly, clear cell carcinoma, which is also considered to originate from endometrial cells (#62), showed a higher EM/FT methylation score than HGSC (Figure2C, P=1.1x10^−34^, Supplementary figure %8%B).

**Figure 2).**
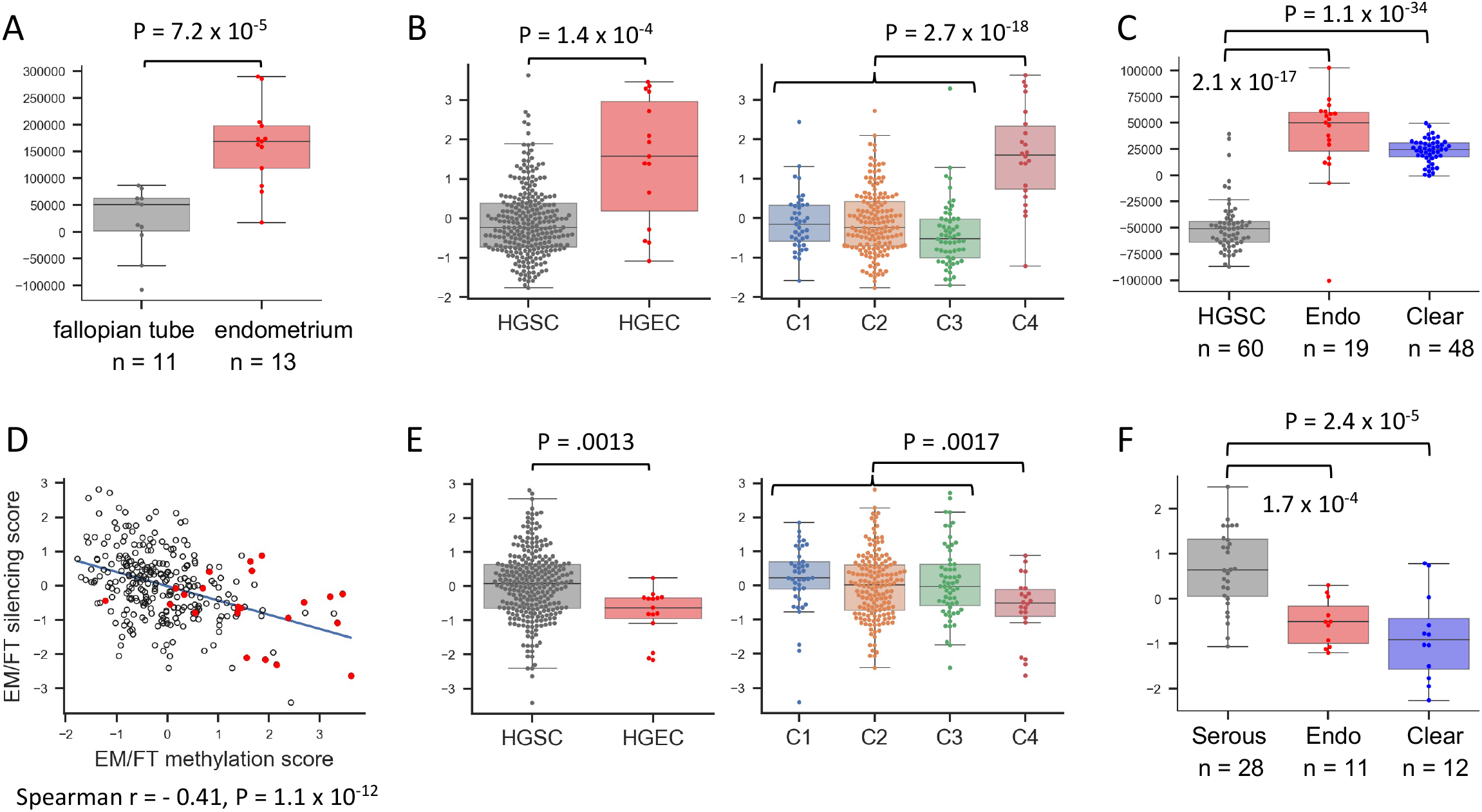
Genome-wide DNA methylation analysis associated with cell differentiation of endometrium and fallopian tubes. A) Comparison of EM/FT methylation score between fallopian tube samples and endometrium samples In GSE155760, EM/FT methylation score was significantly higher in endometrium samples than fallopian tube samples. B) Comparison of EM/FT methylation score in the JGOG-TR2. EM/FT methylation score was significantly higher in tumors diagnosed as HGEC by CPR than those as HGSC (left) and in the C4 tumors than the other tumors (right). C) Comparison of EM/FT methylation score between different histological types of ovarian cancer. In GSE226823, EM/FT methylation score was higher in endometrioid carcinoma (Endo) or in clear cell carcinoma (Clear) than HGSC. D) Correlation of EM/FT methylation score and EM/FT silencing score in the JGOG-TR2 cohort. the EM/FT methylation score and the EM/FT silencing score were negatively correlated. E) Comparison of EM/FT silencing score in the JGOG-TR2. EM/FT silencing score was significantly lower in tumors diagnosis as HGEC than those as HGSC (left) and in the C4 tumors than the other tumors (right). F) Comparison of EM/FT silencing score between different histological types of ovarian cancer. In GSE44104, the EM/FT silencing score was significantly higher in Endo or Clear than in Serous.

From the gene expression profiles of the 208 genes in the EM>FT DMRs and the 106 genes in the FT>EM DMRs, we developed the EM/FT silencing score (see Methods). As expected, the EM/FT silencing score negatively correlated with the EM/FT methylation score in the JGOG-TR2 cohort (Spearman r=-0.41, P=1.1x10^−12^, Figure 2D) and was lower both in the CPR-based HGEC (P=.0013) and C4 (P=.0017) than the other tumors (Figure 2E). In another gene expression dataset for ovarian cancer (GSE44104) (#48), the EM/FT silencing score was consistently lower in endometroid carcinoma and clear cell carcinoma than serous carcinoma (Figure 2F, P=1.7x10^−4^ and 2.4x10^−5^). This result was reproduced in other gene expression datasets (Supplementary figure %8%C).

### Analysis of the TCGA-OV dataset

To investigate whether the TCGA-OV cases include HGEC-type tumors, we built a machine-learning model that classifies tumors into the four aforementioned subtypes based on CNV signatures. We adopted five different classification algorithms, namely, k-nearest neighbor, support vector machine, random forest, multilayer perceptron, and XGBoost, where all of them showed high accuracy of around 95% in the training dataset (Supplementary figure %9%A).

Using the CNV signature of TCGA-OV data as input, after adjusting for batch effects with the JGOG-TR2 cohort (Supplementary figure %9%B), tumor subtypes were determined when four or more of the prediction results from the five classifiers matched, otherwise excluded as undetermined cases.

Subtyping was successful in 509 of 555 cases (92%), with 102 cases predicted to be C1(pC1), 237 to pC2, 158 to pC3, and 13 pC4/HGEC-type tumors. The pC4 showed similar genomic profiles to the C4 in the JGOG-TR2, including lower *TP53* mutation rate (P=3.8x10^−10^), absence of *BRCA1/2* alteration (P=.047), absence of *CCNE1* amplifications (P=.013), higher frequency of *PIK3CA/ARID1A/KRAS/PTEN* mutations (P=5.5x10^−6^), lower HRD score (P=3.2x10^−13^), lower ploidy (P=6.2x10^−4^), and lower MATH score (P=1.9x10^−6^) (Figure 3A). In gene expression, the pC4 showed higher JGOG-C4 signature score (see Methods) (P=8.7x10^−6^), lower KEGG_CELL_CYCLE (P=.0024) score, lower G2M_CHECKPOINT score (P=.014), and lower *WT1* gene expression level (P=1.7x10^−5^) compared to the other groups (Figure 3A). The EM/FT silencing gene score was also lower in the pC4 (P=.0032, Figure 3A).

**Figure 3).**
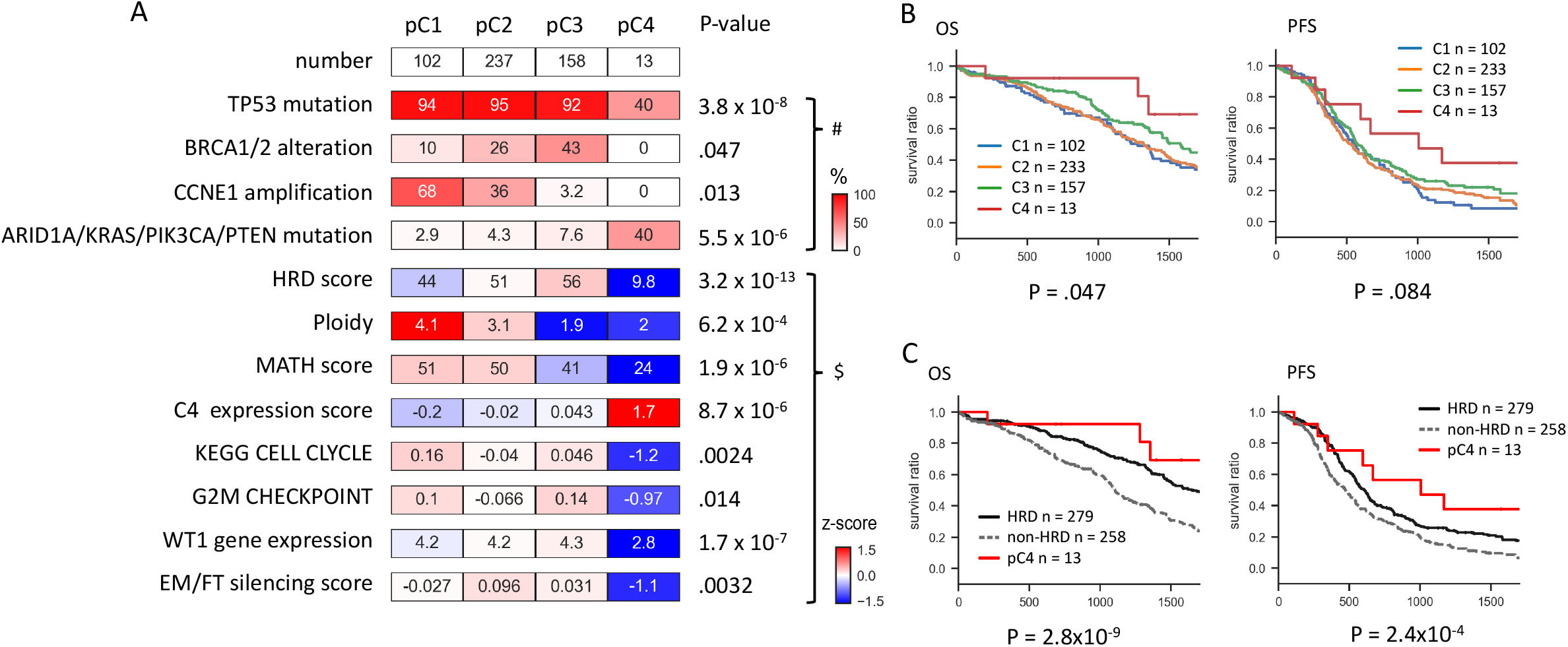
Analysis in TCGA-OV datasets. A) Differences in genomic profiles between the predicted tumor subtypes. Observed differences in molecular profiles between the assigned groups were similar to those in the JGOG-TR2 cohort (Figure1A, Supplementary figure%7%A). Binary (#) and continuous ($) values were compared between C4 and other subtypes using Chi-square test and T-test, respectively. B) Differences of OS (left) and PFS (right) outcomes between the predicted four subtypes. C) Differences in OS (left) and PFS (right) outcomes between HRD, pC4, and non-HRD without pC4.

Survival analysis showed that the pC4 cases tended to have better OS (P=.047) and PFS (P=.083) (Figure 3B) among the four subtypes. Notably, even though all pC4 cases were classified as non-HRD based on *BRCA* alterations and HRD score (see Methods), they tended to have better OS and PFS than other non-HRD cases (Figure 3C).

These findings indicate that a small proportion (2.6%; 13/509) of HGEC-type tumors are included even in the TCGA-OV cohort, where only tumors diagnosed as HGSC were enrolled.

### Analysis of the TCGA-UCEC dataset

Similar to ovarian cancer, uterine endometrial cancer includes both grade 3 endometrioid carcinoma (UHGEC) and serous carcinoma (USEC) histology. The same prediction method was applied to tumors with diagnoses of UHGEC and USEC in the TCGA-UCEC cohort.

Of the total 287 cases, tumor subtypes were successfully determined in 251 (87%) cases, comprising 183 UHGEC and 104 USEC cases. TCGA-UCEC samples predicted as C4 (UpC4) had similar CNV signature profiles to the pC4 of TCGA-OV (Figure 4A) and most of the UpC4 (96%, 112/117) were UHGEC. The genomic profiles of the predicted subtypes were similar to those in JGOG-TR2 and TCGA-OV cohort (Figure 4B); the UpC4 showed lower *TP53* mutation rate (P=7.4x10^−24^), absence of *CCNE1* amplifications (P=1.5x10^−8^), higher frequency of *PIK3CA/ARID1A/KRAS/PTEN* mutations (P=8.5x10^−6^), lower HRD score (P=6.4x10^−56^), lower ploidy (P=3.9x10^−28^), lower MATH score (P=4.8x10^−21^), higher JGOG-C4 signature score (P=1.4x10^−21^), lower *WT1* gene expression (P=9.8x10^−10^), higher EM/FT methylation score (P=1.3x10^−13^), and lower EM/FT silencing score (P=2.0x10^−7^) than the UpC1-UpC3. These differences were significant even after excluding the POLE/dMMR tumors (Supplementary figure %10%A). UpC4 tended to have better survival outcomes than the others (Supplementary figure %10%B), but there was no significant difference among the four subtypes after excluding POLE/dMMR tumors (Supplementary figure %10%C).

**Figure 4).**
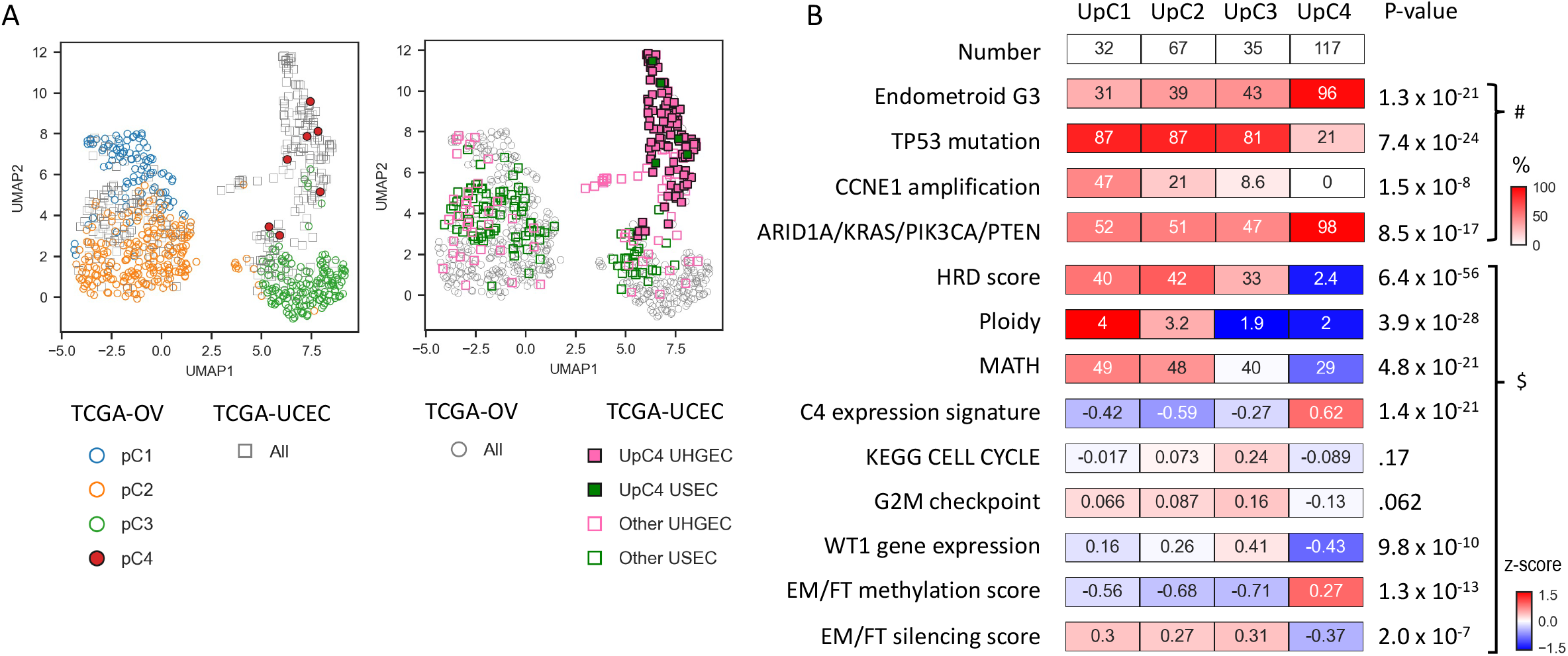
Analysis in TCGA-UCEC. A) Sample distribution from TCGA-OV and TCGA-UCEC cases using UMAP plots in relation to predicted subtype and histology. (left) pC1, pC2, pC3 and pC4 tumors in TCGA-OV were denoted by blue rings, orange rings, green rings, and red solid dots, respectively. TCGA-UCEC tumors were represented by grey square frames. (right) High-grade endometroid (UHGEC) and serous carcinoma (USEC) in TCGA-UCEC were colored in pink and green, and UpC4 and the other tumors were depicted as solid squares and square frames, respectively. TCGA-OV tumors were represented by grey rings. B) Differences in genomic profiles between predicted tumor subtypes. Observed differences in molecular profiles between the assigned groups were similar to those in the JGOG-TR2 cohort (Figure1A, Supplementary figure%7%A). Binary (#) and continuous ($) values were compared between C4 and other subtypes using Chi-square test and T-test, respectively.

In another DNA methylation array dataset for uterine endometrial cancer (GSE155760), the EM/FT methylation score was higher in endometrioid carcinoma than in serous carcinoma (P=1.6 x10^−4^, Supplementary figure %11%A). And in another gene expression dataset for uterine endometrial cancer (GSE23528), the EM/FT silencing score was lower in endometrioid carcinoma than in serous carcinoma (P=.027, Supplementary figure %11%B).

## Discussion

To our knowledge, JGOG3025-TR2 is a unique multi-omics dataset that includes both HGSCs and HGECs of the ovary. In this study, we comprehensively analyzed this dataset and identified a group of tumors with characteristic molecular profiles that differ from HGSCs, which we defined as “HGEC-type” tumors. The HGEC-type tumors were found in a subset of tumors morphologically diagnosed as HGSCs (Figure 1A, 3A).

The low *WT1* mRNA expression in C4 (Figure 1A) and pC4 (Figure 3A) indicate that those tumors are closer to HGEC than HGSC (#57, #63, #64, Supplementary figure %6%). Importantly, even within the C1, C2, and C3 tumors, which were considered as typical HGSCs, some tumors showed low *WT1* gene expression (Figure 1A). It has been reported that about 20% of HGSCs showed negative WT1 expression (#63, #65, #66) and such cases had a poor prognosis in HGSCs (#65). Given the much lower frequency of HGEC relative to HGSC in ovarian cancer, if HGECs are diagnosed based on WT1 negativity, it is more likely to that HGSCs with negative WT1 will be included. In a previous study using whole exome sequencing data of 112 ovarian endometrioid carcinomas (#56), tumors were enrolled based on WT1 negativity. As a result, in addition to eight tumors of grade 3 endometrioid carcinoma, 15 “tumors with serous-like features” were included, and 12 of these tumors had *TP53* mutations. Those tumors could belong to the HGSC (C1,C2,C3) groups according to our CNV signature classification.

There are conflicting reports regarding the proportion of HRD-positive tumors in endometrioid carcinoma (#67, #68). In a single-center retrospective cohort of ovarian endometrioid carcinoma, HRD tumors were found in 19% (5/26) of ovarian endometroid carcinomas (#67). However, in a prospective study of centrally pathologically reviewed epithelial ovarian carcinomas, only three cases of HGEC were observed among 125 cases with *BRCA1/2* pathogenic variants (#68). And of these three, one was WT1-positive and one was *POLE*-mutated, whose *BRCA* mutation may be a passenger variant, suggesting that pathogenic variants in *BRCA1/2* are extremely rare in true endometrioid carcinoma (#68). In the present study, C4/pC4 tumors, or molecularly defined “HGEC-type” tumors, were all non-HRD tumors in terms of *BRCA* alterations and HRD scores (Figure 1A, Supplementary figure %7%, Figure 3A). In general, non-HRD HGSCs have a poor prognosis compared to HRD tumors due to their chemoresistance (#25, #69). However, HGEC-type tumors, despite being non-HRD tumors, exhibited relatively favorable outcomes (Figure 1C, Figure 3C). Since tumor aneuploidy (#70), tumor heterogeneity (#71), and tumor proliferation rate (#72, #73) are associated with poor prognosis, lower scores on these factors (Figure 1A, Supplementary figure %7%, Figure 3A) may be attributed to their favorable outcomes. On the other hand, a substantial proportion of HGEC-type tumors had gene mutations in *ARID1A, PIK3CA, KRAS, and PTEN* (Figure S2A, Figure 3A), similar to low-grade endometrioid carcinomas (#55, #74). This suggests that these tumors may benefit from targeted therapies that target these specific gene mutations rather than PARP inhibitors (#75).

The cell of origin for epithelial ovarian carcinoma is thought to differ among histological types; HGSC arises from serous tubal intraepithelial carcinoma (STIC) originating from fallopian tubal epithelial cells (#76), while endometrioid carcinoma and clear cell carcinoma arise from (ectopic) endometrial cells retrogradely implanted onto the ovary from the uterine endometrium (#2, #75, #77). We found that the differences in methylation status between normal fallopian tube cells and endometrial cells are replicated between ovarian HGSCs and endometrioid or clear cell carcinomas (Figure 2). The finding that C4 tumors in JGOG-TR2 and pC4 tumors in TCGA-OV showed methylation profiles similar to endometrial cells (Figure 2, Figure 3A) strongly suggests that they are of endometrial origin (#78, #79). Here, we utilized whole-genome bisulfite sequencing (WGBS) (#36) data as a training set to identify differentially methylated regions (Supplementary figure %8%A). Non-cancerous cells generally have less DNA methylation in CpG islands compared to cancer cells. The use of WGBS data, which is highly sensitive and comprehensive, was optimal for comparative analysis of methylation status between normal cells.

Interestingly, most of the high-grade carcinomas in TCGA-UCEC were successfully classified into the four subtypes similar to those in ovarian cancer based on CNV signatures (Figure 4). Since UpC1, UpC2, and UpC3 tumors showed methylation status for tubal cell differentiation and UpC4 for endometrial cell differentiation, the difference between them could be attributed to differences in cell of origin. The precursor lesion of serous endometrial cancer has traditionally been considered serous endometrial intraepithelial carcinoma (SEIC), but it is also possible that tumor cells derived from STIC detach from the fallopian tube, migrate into the uterine cavity, and subsequently implant in the endometrium to form a tumor. Recently, it has been reported that tumor cells derived from STIC have been identified in cervical cytology specimens obtained before the diagnosis of ovarian HGSC (#80). There is an increased risk of serous endometrial cancer after menopause (#81), when intermittent shedding of the endometrium ceases. It has been reported that 20% of serous endometrial cancers are associated with STIC-like tubal lesions (#82) and that SEIC and STIC often coexist (#83). Germline carriers of *BRCA1/2* mutations are at risk for serous endometrial cancer (#84). These observations are consistent with our hypothesis. To validate this hypothesis, further research is warranted.

A limitation of this study is the relatively small number of cases molecularly diagnosed as C4/HGEC-type (n=22) in JGOG-TR2. Secondly, the 51 genes targeted for DNA sequencing in this study were primarily selected based on the association with the homologous recombination pathway, and *CTNNB1*, which is frequently mutated in endometrioid carcinoma, was not included. Thirdly, tumors with MSI-high or TMB-high were examined from a relatively small genomic region (approximately 180KB), potentially leading to inaccurate annotations. Finally, it may not be practical to apply SNP array-based CNV signature analysis to clinicopathological diagnosis. Recently, CNV analysis has increasingly been performed using whole genome sequencing, which allows more comprehensive analysis of single nucleotide variants, small insertions and deletions, and structural variants (#85). Future comprehensive genomic analysis of a larger number of cases will reveal the genomic profile of HGEC in more detail.

In conclusion, we identified CNV signature profiles characterizing both HGEC and HGSC using the JGOG-TR2 multi-omics analysis cohort. This study not only elucidated the nature of HGEC but also revealed the presence of molecularly diagnosed HGEC-type tumors within tumors diagnosed as HGSC. The findings will provide new insights into the pathological diagnosis of ovarian cancer and facilitate individualized treatment approaches in the future.

**Table 1.**

| Histology |  | High-grade endometrioid<br>(HGEC) | High-grade serous<br>(HGSC) |
| --- | --- | --- | --- |
| Number of patient |  | 15 | 274 |
| Stage | II | 2 (13%) | 26 (9%) |
|  | III | 6 (40%) | 189 (69%) |
|  | IV | 7 (47%) | 53 (19%) |
|  | Unknown |  | 6 (2%) |
| Age | Median [25<sup>th</sup>, 75<sup>th</sup>] | 59 [53, 68] | 61.5 [54, 69] |
| Performance status | 0 | 12 (80%) | 202 (74%) |
|  | >=1 | 2 (13%) | 64 (23%) |
|  | Unknown | 1 (7%) | 8 (3%) |
| Residual tumor | 0 | 10 (67%) | 161 (59%) |
|  | 1cm | 3 (20%) | 44 (16%) |
|  | >= 1cm | 2 (13%) | 69 (25%) |
| Follow-up day | Median [25<sup>th</sup>, 75<sup>th</sup> ] | 1310 [915, 1399] | 1072 [932, 1229] |
| Analysis type | Targeted DNA sequencing | 15 | 274 |
|  | RNA sequencing | 15 | 271 |
|  | DNA methylation array | 15 | 268 |
|  | SNP array | 15 | 272 |

## Data Availability

Processed data and analysis codes to reproduce the results in this study are available in the GitHub project page (https://github.com/shirotak/JGOG_HGEOC). Genomic data of the JGOG3025-TR2 cohort, including RNA-seq, SNP array, and DNA methylation array are available from SRA (PRJNA1092599) and NCBI-GEO (GSE263455). Targeted sequencing data and clinical information are available upon reasonable request to JGOG (, https://jgog.gr.jp/en/index.html).

https://www.ncbi.nlm.nih.gov/geo/query/acc.cgi?acc=GSE263455

## Acknowledgements

This work was supported by AstraZeneca K.K. and Merck Sharp & Dohme Corp as the programme of Externally Sponsored Research (ESR-19-14550) and in part by the Japan Agency for Medical Research and development, AMED under Grant Number JP21tm0124005. We would like to thank all the JGOG members who participated in the JGOG3025-TR2 study; Drs. Makio Shozu, Hiroyuki Shigeta, Kazuhiro Takehara, Akira Kikuchi, Toyomi Sato, Akinori Oki, Shinya Yoshioka, Shinya Sato, Ryuji Kawaguchi, Hisafumi Okura, Takeshi Iwasa, Shoji Kamiura, Masato Kamitomo, Yoichi Aoki, Nao Suzuki, Yoshio Yoshida, Tadashi Kimura, Daisuke Aoki, Kazuyoshi Kato, Hiroaki Kobayashi, Hidemichi Watari, Etsuko Miyagi, Tsuyoshi Saito, Yoshihito Yokoyama, Tsunekazu Kita, Takashi Matsumoto, Satoshi Nagase, Toshiya Yamamoto, Yukio Hirano, Tomoaki Ikeda, Shiro Suzuki, Keiya Fujimori, Nagamasa Maeda, Naohiko Umesaki, Masatoshi Sugita, and Akira Kouyama.

## Author contributions

ST: data analysis and writing the manuscript; RTH: data analysis and review of the manuscript ; KY: sample collection and data analysis; TB: sample collection and review of the manuscript; MS: sample collection and review of the manuscript; HY: sample collection and review of the manuscript; AO: sample collection and review of the manuscript; HK: sample collection and review of the manuscript; KO: sample collection and data analysis; MM: data analysis and review of the manuscript; TE: sample collection and review of the manuscript; NM: design of this study, data analysis and writing the manuscript.

## Ethics

For the JGOG3025 study, i.e., clinical data analysis, frozen tumour tissue collection, target sequencing, and future analyses of tumour tissue, written informed consent was obtained from all patients with approval from the Institutional Review Board at each JGOG participating site prior to the start of the study (#7). The JGOG3025-TR2 study was then conducted with the approval of the Ethics Committee of JGOG and the Institutional Ethics Committee of Kindai University (approval number; 29-167), with opt-out patient consent. This study was performed in accordance with the Declaration of Helsinki.

## References

1. PMID:31640608 Zhang, Y., Luo, G., Li, M., Guo, P., Xiao, Y., Ji, H., & Hao, Y. (2019). Global patterns and trends in ovarian cancer incidence: age, period and birth cohort analysis. BMC Cancer, 19(1), 984. doi:10.1186/s12885-019-6139-6

2. PMID:31640608 Nakai, H., Higashi, T., Kakuwa, T., & Matsumura, N.(in press). Trends in gynecologic cancer in Japan: incidence from 1980 to 2019 and mortality from 1981 to 2021. International journal of clinical oncology. doi:10.1007/s10147-024-02473-8

3. PMID:31640608 Koshiyama, M., Matsumura, N., & Konishi, I. (2014). Recent concepts of ovarian carcinogenesis: type I and type II. BioMed Research International, 2014, 934261. doi:10.1155/2014/934261

4. PMID:31640608 Yamaguchi, K., Mandai, M., Oura, T., Matsumura, N., Hamanishi, J., Baba, T., … Konishi, I. (2010). Identification of an ovarian clear cell carcinoma gene signature that reflects inherent disease biology and the carcinogenic processes. Oncogene, 29(12), 1741–1752. doi:10.1038/onc.2009.470

5. PMID:31640608 Yamaguchi, Ken, Huang, Z., Matsumura, N., Mandai, M., Okamoto, T., Baba, T., … Murphy, S. K. (2014). Epigenetic determinants of ovarian clear cell carcinoma biology. International Journal of Cancer. Journal International Du Cancer, 135(3), 585–597. doi:10.1002/ijc.28701

6. PMID:31640608 Huang, R. Y., Chen, G. B., Matsumura, N., Lai, H.-C., Mori, S., Li, J., … Goh, L. (2012). Histotype-specific copy-number alterations in ovarian cancer. BMC Medical Genomics, 5(1), 47. doi:10.1186/1755-8794-5-47

7. PMID:31640608 Yoshihara, K., Baba, T., Tokunaga, H., Nishino, K., Sekine, M., Takamatsu, S., … Enomoto, T. (2023). Homologous recombination inquiry through ovarian malignancy investigations: JGOG3025 Study. Cancer Science, 114(6), 2515–2523. doi:10.1111/cas.15747

8. PMID:31640608 Nakai, H., & Matsumura, N. (2022b). The roles and limitations of bevacizumab in the treatment of ovarian cancer. International Journal of Clinical Oncology, 27(7), 1120–1126. doi:10.1007/s10147-022-02169-x

9. PMID:31640608 Nakai, H., & Matsumura, N. (2022a). Individualization in the first-line treatment of advanced ovarian cancer based on the mechanism of action of molecularly targeted drugs. International Journal of Clinical Oncology, 27(6), 1001–1012. doi:10.1007/s10147-022-02163-3

10. PMID:31640608 Jang, J. Y. A., Yanaihara, N., Pujade-Lauraine, E., Mikami, Y., Oda, K., Bookman, M., … Okamoto, A. (2017). Update on rare epithelial ovarian cancers: based on the Rare Ovarian Tumors Young Investigator Conference. Journal of Gynecologic Oncology, 28(4), e54. doi:10.3802/jgo.2017.28.e54

11. PMID:31640608 Mandai, M., Yamaguchi, K., Matsumura, N., Baba, T., & Konishi, I. (2009). Ovarian cancer in endometriosis: molecular biology, pathology, and clinical management. International Journal of Clinical Oncology, 14(5), 383–391. doi:10.1007/s10147-009-0935-y

12. PMID:31640608 Soyama, H., Miyamoto, M., Takano, M., Iwahashi, H., Kato, K., Sakamoto, T., … Furuya, K. (2018). A pathological study using 2014 WHO criteria reveals poor prognosis of grade 3 ovarian endometrioid carcinomas. In Vivo (Athens, Greece), 32(3), 597–602. doi:10.21873/invivo.11281

13. PMID:31640608 Schwartz, D. R., Kardia, S. L. R., Shedden, K. A., Kuick, R., Michailidis, G., Taylor, J. M. G., … Cho, K. R. (2002). Gene expression in ovarian cancer reflects both morphology and biological behavior, distinguishing clear cell from other poor-prognosis ovarian carcinomas. Cancer Research, 62(16), 4722–4729. Retrieved from https://www.ncbi.nlm.nih.gov/pubmed/12183431

14. PMID:31640608 Tothill, R. W., Tinker, A. V., George, J., Brown, R., Fox, S. B., Lade, S., … Bowtell, D. D. L. (2008). Novel molecular subtypes of serous and endometrioid ovarian cancer linked to clinical outcome. Clinical Cancer Research: An Official Journal of the American Association for Cancer Research, 14(16), 5198–5208. doi:10.1158/1078-0432.CCR-08-0196

15. WHO2014) WHO classification of tumours of female reproductive organs. 4th Edicion, Volume 6. IARC Press; 2014.

16. WHO2020) WHO classification of tumours of female reproductive organs. 5th Edition, Volume 4. IARC Press; 2020.

17. PMID:31640608 Lim, D., Murali, R., Murray, M. P., Veras, E., Park, K. J., & Soslow, R. A. (2016). Morphological and immunohistochemical reevaluation of tumors initially diagnosed as ovarian endometrioid carcinoma with emphasis on high-grade tumors. The American Journal of Surgical Pathology, 40(3), 302–312. doi:10.1097/PAS.0000000000000550

18. PMID:31640608 Mirza, M. R., Monk, B. J., Herrstedt, J., Oza, A. M., Mahner, S., Redondo, A., … ENGOT-OV16/NOVA Investigators. (2016). Niraparib maintenance therapy in platinum-sensitive, recurrent ovarian cancer. The New England Journal of Medicine, 375(22), 2154–2164. doi:10.1056/NEJMoa1611310

19. PMID:31640608 Ledermann, J., Harter, P., Gourley, C., Friedlander, M., Vergote, I., Rustin, G., … Matulonis, U. (2012). Olaparib maintenance therapy in platinum-sensitive relapsed ovarian cancer. The New England Journal of Medicine, 366(15), 1382–1392. doi:10.1056/NEJMoa1105535

20. PMID:31640608 Moore, K., Colombo, N., Scambia, G., Kim, B.-G., Oaknin, A., Friedlander, M., … DiSilvestro, P. (2018). Maintenance olaparib in patients with newly diagnosed advanced ovarian cancer. The New England Journal of Medicine, 379(26), 2495–2505. doi:10.1056/NEJMoa1810858

21. PMID:31640608 Ray-Coquard, I., Pautier, P., Pignata, S., Pérol, D., González-Martín, A., Berger, R., … PAOLA-1 Investigators. (2019). Olaparib plus bevacizumab as first-line maintenance in ovarian cancer. The New England Journal of Medicine, 381(25), 2416–2428. doi:10.1056/NEJMoa1911361

22. PMID:31640608 González-Martín, A., Pothuri, B., Vergote, I., DePont Christensen, R., Graybill, W., Mirza, M. R., … PRIMA/ENGOT-OV26/GOG-3012 Investigators. (2019). Niraparib in patients with newly diagnosed advanced ovarian cancer. The New England Journal of Medicine, 381(25), 2391–2402. doi:10.1056/NEJMoa1910962

23. PMID:31640608 Li, N., Zhu, J., Yin, R., Wang, J., Pan, L., Kong, B., … Wu, L. (2023). Treatment with niraparib maintenance therapy in patients with newly diagnosed advanced ovarian cancer: A phase 3 randomized clinical trial. JAMA Oncology, 9(9), 1230–1237. doi:10.1001/jamaoncol.2023.2283

24. PMID:31640608 De’ Angelis, G. L., Bottarelli, L., Azzoni, C., De’ Angelis, N., Leandro, G., Di Mario, F., … Negri, F. (2018). Microsatellite instability in colorectal cancer. Acta Bio-Medica : Atenei Parmensis, 89(9-S), 97–101. doi:10.23750/abm.v89i9-S.7960

25. PMID:31640608 Takamatsu, S., Yoshihara, K., Baba, T., Shimada, M., Yoshida, H., Kajiyama, H., … Matsumura, N. (2023). Prognostic relevance of HRDness gene expression signature in ovarian high-grade serous carcinoma; JGOG3025-TR2 study. British Journal of Cancer, 128(6), 1095–1104. doi:10.1038/s41416-022-02122-9

26. PMID:31640608 Wang, K., Li, M., & Hakonarson, H. (2010). ANNOVAR: functional annotation of genetic variants from high-throughput sequencing data. Nucleic Acids Research, 38(16), e164. doi:10.1093/nar/gkq603

27. PMID:31640608 Chakravarty, D., Gao, J., Phillips, S. M., Kundra, R., Zhang, H., Wang, J., … Schultz, N. (2017). OncoKB: A precision oncology knowledge base. JCO Precision Oncology, 2017(1), 1–16. doi:10.1200/PO.17.00011

28. PMID:31640608 Cline, M. S., Liao, R. G., Parsons, M. T., Paten, B., Alquaddoomi, F., Antoniou, A., … Spurdle, A. B. (2018). BRCA Challenge: BRCA Exchange as a global resource for variants in BRCA1 and BRCA2. PLoS Genetics, 14(12), e1007752. doi:10.1371/journal.pgen.1007752

29. PMID:31640608 Van Loo, P., Nordgard, S. H., Lingjærde, O. C., Russnes, H. G., Rye, I. H., Sun, W., … Kristensen, V. N. (2010). Allele-specific copy number analysis of tumors. Proceedings of the National Academy of Sciences of the United States of America, 107(39), 16910–16915. doi:10.1073/pnas.1009843107

30. PMID:31640608 Khandekar, A., Vangara, R., Barnes, M., Díaz-Gay, M., Abbasi, A., Bergstrom, E. N., … Alexandrov, L. B. (2023). Visualizing and exploring patterns of large mutational events with SigProfilerMatrixGenerator. BMC Genomics, 24(1), 469. doi:10.1186/s12864-023-09584-y

31. PMID:31640608 Sztupinszki, Z., Diossy, M., Krzystanek, M., Reiniger, L., Csabai, I., Favero, F., … Szallasi, Z. (2018). Migrating the SNP array-based homologous recombination deficiency measures to next generation sequencing data of breast cancer. NPJ Breast Cancer, 4, 16. doi:10.1038/s41523-018-0066-6

32. PMID:31640608 Ritchie, M. E., Phipson, B., Wu, D., Hu, Y., Law, C. W., Shi, W., & Smyth, G. K. (2015). limma powers differential expression analyses for RNA-sequencing and microarray studies. Nucleic Acids Research, 43(7), e47. doi:10.1093/nar/gkv007

33. PMID:31640608 Law, C. W., Chen, Y., Shi, W., & Smyth, G. K. (2014). voom: Precision weights unlock linear model analysis tools for RNA-seq read counts. Genome Biology, 15(2), R29. doi:10.1186/gb-2014-15-2-r29

34. PMID:31640608 Love, M. I., Huber, W., & Anders, S. (2014). Moderated estimation of fold change and dispersion for RNA-seq data with DESeq2. Genome Biology, 15(12), 550. doi:10.1186/s13059-014-0550-8

35. PMID:31640608 Barbie, D. A., Tamayo, P., Boehm, J. S., Kim, S. Y., Moody, S. E., Dunn, I. F., … Hahn, W. C. (2009). Systematic RNA interference reveals that oncogenic KRAS-driven cancers require TBK1. Nature, 462(7269), 108–112. doi:10.1038/nature08460

36. PMID:31640608 Loyfer, N., Magenheim, J., Peretz, A., Cann, G., Bredno, J., Klochendler, A., … Kaplan, T. (2023). A DNA methylation atlas of normal human cell types. Nature, 613(7943), 355–364. doi:10.1038/s41586-022-05580-6

37. PMID:31640608 Minegishi, R., Gotoh, O., Tanaka, N., Maruyama, R., Chang, J. T., & Mori, S. (2021). A method of sample-wise region-set enrichment analysis for DNA methylomics. Epigenomics, 13(14), 1081–1093. doi:10.2217/epi-2021-0065

38. PMID:31640608 Cavalcante, R. G., & Sartor, M. A. (2017). Annotatr: Genomic regions in context. Bioinformatics (Oxford, England), 33(15), 2381–2383. doi:10.1093/bioinformatics/btx183

39. PMID:31640608 Cancer Genome Atlas Research Network. (2011). Integrated genomic analyses of ovarian carcinoma. Nature, 474(7353), 609–615. doi:10.1038/nature10166

40. PMID:31640608 Cancer Genome Atlas Research Network, Kandoth, C., Schultz, N., Cherniack, A. D., Akbani, R., Liu, Y., … Levine, D. A. (2013). Integrated genomic characterization of endometrial carcinoma. Nature, 497(7447), 67–73. doi:10.1038/nature12113

41. PMID:31640608 Grossman, R. L., Heath, A. P., Ferretti, V., Varmus, H. E., Lowy, D. R., Kibbe, W. A., & Staudt, L. M. (2016). Toward a shared vision for cancer genomic data. The New England Journal of Medicine, 375(12), 1109–1112. doi:10.1056/NEJMp1607591

42. PMID:31640608 Bartlett, T. E., Jones, A., Goode, E. L., Fridley, B. L., Cunningham, J. M., Berns, E. M. J. J., … Widschwendter, M. (2015). Intra-gene DNA methylation variability is a clinically independent prognostic marker in women’s cancers. PloS One, 10(12), e0143178. doi:10.1371/journal.pone.0143178

43. PMID:31640608 Pisanic, T. R., 2nd, Wang, Y., Sun, H., Considine, M., Li, L., Wang, T.-H., … Shih, I.-M. (2020). Methylomic landscapes of ovarian cancer precursor lesions. Clinical Cancer Research: An Official Journal of the American Association for Cancer Research, 26(23), 6310–6320. doi:10.1158/1078-0432.CCR-20-0270

44. PMID: 31640608 Chan, David W et al. “Genome-wide DNA methylome analysis identifies methylation signatures associated with survival and drug resistance of ovarian cancers.” Clinical epigenetics vol. 13,1 142. 22 Jul. 2021, doi:10.1186/s13148-021-01130-5

45. PMID:31640608 Beddows, I., Fan, H., Heinze, K., Johnson, B. K., Leonova, A., Senz, J., … Shen, H. (2024). Cell state of origin impacts development of distinct endometriosis-related ovarian carcinoma histotypes. Cancer Research, 84(1), 26–38. doi:10.1158/0008-5472.CAN-23-1362

46. PMID: 31640608 Wu, Rong et al. “Impact of oviductal versus ovarian epithelial cell of origin on ovarian endometrioid carcinoma phenotype in the mouse.” The Journal of pathology vol. 240,3 (2016): 341–351. doi:10.1002/path.4783

47. PMID: 31640608 Ramakrishna, Manasa et al. “Identification of candidate growth promoting genes in ovarian cancer through integrated copy number and expression analysis.” PloS one vol. 5,4 e9983. 8 Apr. 2010, doi:10.1371/journal.pone.0009983

48. PMID: 31640608 Wu, Y-H et al. “COL11A1 promotes tumor progression and predicts poor clinical outcome in ovarian cancer.” Oncogene vol. 33,26 (2014): 3432–40. doi:10.1038/onc.2013.307

49. PMID: 31640608 Uehara, Yuriko et al. “Correction: Integrated Copy Number and Expression Analysis Identifies Profiles of Whole-Arm Chromosomal Alterations and Subgroups with Favorable Outcome in Ovarian Clear Cell Carcinomas.” PloS one vol. 10,7 e0132751. 6 Jul. 2015, doi:10.1371/journal.pone.0132751

50. PMID:31640608 Desbois, Mélanie et al. “Integrated digital pathology and transcriptome analysis identifies molecular mediators of T-cell exclusion in ovarian cancer.” Nature communications vol. 11,1 5583. 4 Nov. 2020, doi:10.1038/s41467-020-19408-2

51. PMID:31640608 Wu, P., Heins, Z. J., Muller, J. T., Katsnelson, L., de Bruijn, I., Abeshouse, A. A., … Gao, J. (2019). Integration and analysis of CPTAC proteomics data in the context of cancer genomics in the cBioPortal. Molecular & Cellular Proteomics: MCP, 18(9), 1893–1898. doi:10.1074/mcp.TIR119.001673

52. PMID:31640608 Gao, J., Aksoy, B. A., Dogrusoz, U., Dresdner, G., Gross, B., Sumer, S. O., … Schultz, N. (2013). Integrative analysis of complex cancer genomics and clinical profiles using the cBioPortal. Science Signaling, 6(269), 11. doi:10.1126/scisignal.2004088

53. PMID:31640608 Lin, Q., Wagner, W., & Zenke, M. (2013). Analysis of genome-wide DNA methylation profiles by BeadChip technology. Methods in Molecular Biology (Clifton, N.J.), 1049, 21–33. doi:10.1007/978-1-62703-547-7_3

54. PMID:31640608 Zhang, Y., Parmigiani, G., & Johnson, W. E. (2020). ComBat-seq: batch effect adjustment for RNA-seq count data. NAR Genomics and Bioinformatics, 2(3), lqaa078. doi:10.1093/nargab/lqaa078

55. PMID:31640608 Cybulska, P., Paula, A. D. C., Tseng, J., Leitao, M. M., Jr, Bashashati, A., Huntsman, D. G., … Weigelt, B. (2019). Molecular profiling and molecular classification of endometrioid ovarian carcinomas. Gynecologic Oncology, 154(3), 516–523. doi:10.1016/j.ygyno.2019.07.012

56. PMID:31640608 Hollis, R. L., Thomson, J. P., Stanley, B., Churchman, M., Meynert, A. M., Rye, T., … Herrington, C. S. (2020). Molecular stratification of endometrioid ovarian carcinoma predicts clinical outcome. Nature Communications, 11(1), 4995. doi:10.1038/s41467-020-18819-5

57. PMID:31640608 Assem, H., Rambau, P. F., Lee, S., Ogilvie, T., Sienko, A., Kelemen, L. E., & Köbel, M. (2018). High-grade endometrioid carcinoma of the ovary: A clinicopathologic study of 30 cases. The American Journal of Surgical Pathology, 42(4), 534–544. doi:10.1097/PAS.0000000000001016

58. PMID:31640608 Konishi, I., Abiko, K., Hayashi, T., Yamanoi, K., Murakami, R., Yamaguchi, K., … Kyoto Study Group for Ovarian Cancer Research. (2022). Peritoneal dissemination of high-grade serous ovarian cancer: pivotal roles of chromosomal instability and epigenetic dynamics. Journal of Gynecologic Oncology, 33(5), e83. doi:10.3802/jgo.2022.33.e83

59. PMID:31640608 Tao, Z., Wang, S., Wu, C., Wu, T., Zhao, X., Ning, W., … Liu, X.-S. (2023). The repertoire of copy number alteration signatures in human cancer. Briefings in Bioinformatics, 24(2). doi:10.1093/bib/bbad053

60. PMID:31640608 Kuhn, E., Wang, T.-L., Doberstein, K., Bahadirli-Talbott, A., Ayhan, A., Sehdev, A. S., … Shih, I.-M. (2016). CCNE1 amplification and centrosome number abnormality in serous tubal intraepithelial carcinoma: further evidence supporting its role as a precursor of ovarian high-grade serous carcinoma. Modern Pathology: An Official Journal of the United States and Canadian Academy of Pathology, Inc, 29(10), 1254–1261. doi:10.1038/modpathol.2016.101

61. PMID:31640608 Otsuka, I. (2021). Mechanisms of high-grade serous carcinogenesis in the Fallopian tube and ovary: Current hypotheses, etiologic factors, and molecular alterations. International Journal of Molecular Sciences, 22(9). doi:10.3390/ijms22094409

62. PMID:31640608 Cochrane, D. R., Tessier-Cloutier, B., Lawrence, K. M., Nazeran, T., Karnezis, A. N., Salamanca, C., … Huntsman, D. G. (2017). Clear cell and endometrioid carcinomas: are their differences attributable to distinct cells of origin? The Journal of Pathology, 243(1), 26–36. doi:10.1002/path.4934

63. PMID:31640608 Madore, J., Ren, F., Filali-Mouhim, A., Sanchez, L., Köbel, M., Tonin, P. N., … Mes-Masson, A.-M. (2010). Characterization of the molecular differences between ovarian endometrioid carcinoma and ovarian serous carcinoma. The Journal of Pathology, 220(3), 392–400. doi:10.1002/path.2659

64. PMID:31640608 Liu, Z., Yamanouchi, K., Ohtao, T., Matsumura, S., Seino, M., Shridhar, V., … Kurachi, H. (2014). High levels of Wilms’ tumor 1 (WT1) expression were associated with aggressive clinical features in ovarian cancer. Anticancer Research, 34(5), 2331–2340. Retrieved from https://www.ncbi.nlm.nih.gov/pubmed/24778040

65. PMID:31640608 Köbel, M., Kalloger, S. E., Boyd, N., McKinney, S., Mehl, E., Palmer, C., … Huntsman, D. (2008). Ovarian carcinoma subtypes are different diseases: implications for biomarker studies. PLoS Medicine, 5(12), e232. doi:10.1371/journal.pmed.0050232

66. PMID:31640608 Gilks, C. B., Ionescu, D. N., Kalloger, S. E., Köbel, M., Irving, J., Clarke, B., … Cheryl Brown Ovarian Cancer Outcomes Unit of the British Columbia Cancer Agency. (2008). Tumor cell type can be reproducibly diagnosed and is of independent prognostic significance in patients with maximally debulked ovarian carcinoma. Human Pathology, 39(8), 1239–1251. doi:10.1016/j.humpath.2008.01.003

67. PMID:31640608 Pierson, W. E., Peters, P. N., Chang, M. T., Chen, L.-M., Quigley, D. A., Ashworth, A., & Chapman, J. S. (2020). An integrated molecular profile of endometrioid ovarian cancer. Gynecologic Oncology, 157(1), 55–61. doi:10.1016/j.ygyno.2020.02.011

68. PMID:31640608 Kramer, C., Lanjouw, L., Ruano, D., Ter Elst, A., Santandrea, G., Solleveld-Westerink, N., … Bosse, T. (2023). Causality and functional relevance of BRCA1 and BRCA2 pathogenic variants in non-high-grade serous ovarian carcinomas. The Journal of Pathology. doi:10.1002/path.6218

69. PMID:31640608 Takaya, H., Nakai, H., Takamatsu, S., Mandai, M., & Matsumura, N. (2020). Homologous recombination deficiency status-based classification of high-grade serous ovarian carcinoma. Scientific Reports, 10(1), 2757. doi:10.1038/s41598-020-59671-3

70. PMID:31640608 Derrick, M., Englof, I., Drobyshevsky, A., Luo, K., Yu, L., & Tan, S. (2012). Intrauterine fetal demise can be remote from the inciting insult in an animal model of hypoxia-ischemia. Pediatric Research, 72(2), 154–160. doi:10.1038/pr.2012.65

71. PMID:31640608 Takaya, H., Nakai, H., Sakai, K., Nishio, K., Murakami, K., Mandai, M., & Matsumura, N. (2020). Intratumor heterogeneity and homologous recombination deficiency of high-grade serous ovarian cancer are associated with prognosis and molecular subtype and change in treatment course. Gynecologic Oncology, 156(2), 415–422. doi:10.1016/j.ygyno.2019.11.013

72. PMID:31640608 D’Andrilli, G., Kumar, C., Scambia, G., & Giordano, A. (2004). Cell cycle genes in ovarian cancer: steps toward earlier diagnosis and novel therapies. Clinical Cancer Research: An Official Journal of the American Association for Cancer Research, 10(24), 8132–8141. doi:10.1158/1078-0432.CCR-04-0886

73. PMID:31640608 D’Andrilli, G., Giordano, A., & Bovicelli, A. (2008). Epithelial ovarian cancer: the role of cell cycle genes in the different histotypes. The Open Clinical Cancer Journal, 2(1), 7–12. doi:10.2174/1874189400802010007

74. PMID:31640608 Wu, R., Hendrix-Lucas, N., Kuick, R., Zhai, Y., Schwartz, D. R., Akyol, A., … Cho, K. R. (2007). Mouse model of human ovarian endometrioid adenocarcinoma based on somatic defects in the Wnt/beta-catenin and PI3K/Pten signaling pathways. Cancer Cell, 11(4), 321–333. doi:10.1016/j.ccr.2007.02.016

75. PMID:31640608 Murakami, K., Kotani, Y., Nakai, H., & Matsumura, N. (2020). Endometriosis-associated ovarian cancer: The origin and targeted therapy. Cancers, 12(6), 1676. doi:10.3390/cancers12061676

76. PMID:31640608 Reade, Clare J et al. “The fallopian tube as the origin of high grade serous ovarian cancer: review of a paradigm shift.” Journal of obstetrics and gynaecology Canada : JOGC = Journal d’obstetrique et gynecologie du Canada : JOGC vol. 36,2 (2014): 133–140. doi:10.1016/S1701-2163(15)30659-9

77. PMID: 31640608 Garavaglia, Elisabetta et al. “The origin of endometriosis-associated ovarian cancer from uterine neoplastic lesions.” Medical hypotheses vol. 110 (2018): 80–82. doi:10.1016/j.mehy.2017.11.006

78. PMID:31640608 Moran S, Martínez-Cardús A, Sayols S, et al. Epigenetic profiling to classify cancer of unknown primary: a multicentre, retrospective analysis. Lancet Oncol. 2016;17(10):1386–95.

79. PMID:31640608 Papanicolau-Sengos A, Aldape K. DNA Methylation Profiling: An Emerging Paradigm for Cancer Diagnosis. Annu Rev Pathol. 2022;17:295–321.

80. PMID:31640608 Paracchini, L., Mannarino, L., Romualdi, C., Zadro, R., Beltrame, L., Fuso Nerini, I., … Marchini, S. (2023). Genomic instability analysis in DNA from Papanicolaou test provides proof-of-principle early diagnosis of high-grade serous ovarian cancer. Science Translational Medicine, 15(725), eadi2556. doi:10.1126/scitranslmed.adi2556

81. PMID:31640608 Agarwal, A., Yadav, S., Dusane, R., Menon, S., Rekhi, B., & Deodhar, K. K. (2022). Endometrial serous carcinoma: A retrospective review of histological features & their clinicopathological association with disease-free survival & overall survival. The Indian Journal of Medical Research, 156(1), 83–93. doi:10.4103/ijmr.IJMR_697_20

82. PMID:31640608 Kommoss, F., Faruqi, A., Gilks, C. B., Lamshang Leen, S., Singh, N., Wilkinson, N., & McCluggage, W. G. (2017). Uterine serous carcinomas frequently metastasize to the Fallopian tube and can mimic serous tubal intraepithelial carcinoma. The American Journal of Surgical Pathology, 41(2), 161–170. doi:10.1097/PAS.0000000000000757

83. PMID:31640608 Jarboe, E. A., Miron, A., Carlson, J. W., Hirsch, M. S., Kindelberger, D., Mutter, G. L., … Nucci, M. R. (2009). Coexisting intraepithelial serous carcinomas of the endometrium and fallopian tube: frequency and potential significance. International Journal of Gynecological Pathology: Official Journal of the International Society of Gynecological Pathologists, 28(4), 308–315. doi:10.1097/PGP.0b013e3181934390

84. PMID:31640608 de Jonge, M. M., de Kroon, C. D., Jenner, D. J., Oosting, J., de Hullu, J. A., Mourits, M. J. E., … van Asperen, C. J. (2021). Endometrial cancer risk in women with germline BRCA1 or BRCA2 mutations: Multicenter cohort study. Journal of the National Cancer Institute, 113(9), 1203–1211. doi:10.1093/jnci/djab036

85. PMID:31640608 Cortés-Ciriano, Isidro et al. “Computational analysis of cancer genome sequencing data.” Nature reviews. Genetics vol. 23,5 (2022): 298–314. doi:10.1038/s41576-021-00431-y

