## Supplementary data for "Identification of ovarian high-grade endometrioid-type tumors through multi-omics analysis: JGOG3025-TR2 study"

### Contents

### Supplementary figures

Supplementary figure 1) Determination of a cutoff to identify MSI-high tumors in the JGOG3025 whole cohort.

Supplementary figure 2) Determination of the number of clusters

Supplementary figure 3) Differentially expressed gene analysis between C4 vs C1-C3 in the JGOG-TR2

Supplementary figure 4) Trend in percentage of high-grade endometrioid carcinomas among all endometrioid carcinomas of the ovary from 2000 to 2020 in the SEER database

Supplementary figure 5) Differences in genomic profiles between HGEC and HGSC based on CPR diagnosis

Supplementary figure 6) Correlation between protein expression and mRNA expression of in TCGA-OV

Supplementary figure 7) Differences in molecular profiles between the four subtypes in the JGOG-TR2 cohort

Supplementary figure 8) Genome-wide DNA methylation analysis associated with cell differentiation of endometrium and fallopian tubes

Supplementary figure 9) Machine learning-based tumor subtype prediction based on CNV signatures

Supplementary figure 10) Analysis in TCGA-UCEC cohort

Supplementary figure 11) Genome-wide DNA methylation status associated with cell differentiation of endometrium and fallopian tubes in endometrial cancer

### Supplementary tables

Supplementary table 1) JGOG C4 signature UP (147) and DN (161) genes

Supplementary table 2) EM>FT (208) and FT>EM (106) methylation silencing genes

Supplementary table 3) List of publicly available datasets

**Supplementary figure 1) Determination of a cutoff to identify MSI-high tumors in the JGOG3025 whole cohort**

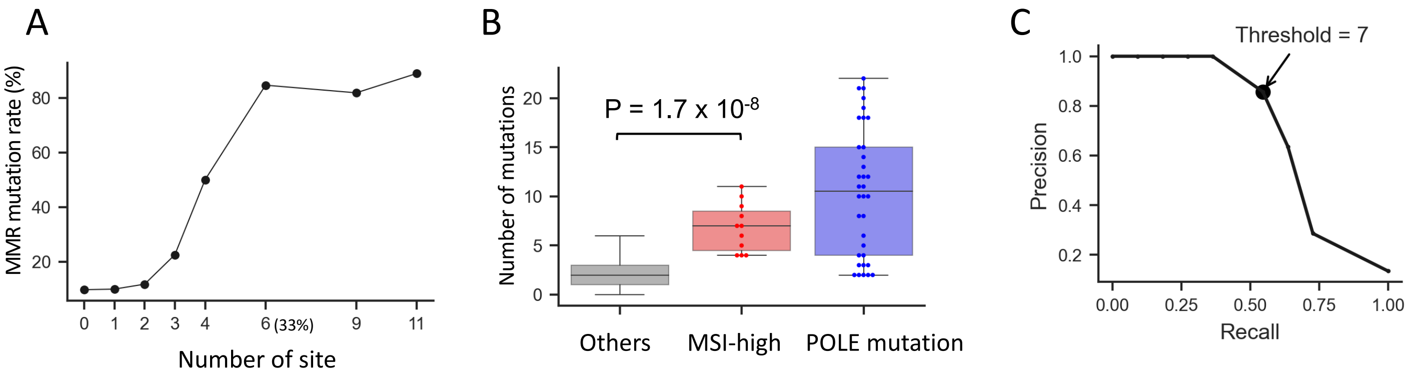


A) Mutation rate of MMR genes in samples with different number of microsatellite sites.
Using MSIsensor2, 18 microsatellite sites were identified in the target DNA sequencing panel used in the JGOG3025 study. The mutation rate in DNA mismatch repair genes including *MLH1, MSH2, MSH6 and PMS2* was >= 80% in samples with 6 or more micro satellite sites. This cutoff is consistent with the previously reported threshold of 30% (De' Angelis GL et al. *Acta Biomed* 2018, PMID: 30561401)

B) The number of detected gene mutations was significantly higher in MSI-high tumors than in the others, except for those with *POLE* mutations.

C) A precision recall curve analysis of the optimal cut-off values for MSI-high tumors.
Excluding *POLE* mutated tumors, a precision recall curve analysis of the number of mutations that most discriminate MSI-high tumors yielded an optimal threshold of 7. Using this cutoff, 29/710 (4.1%) were determined to be TMB-high, which was consistent with the rate of 12/293 (4.1%) in ovarian cancer in a previous study, where tumors with >=175 mutations per whole exome sequencing was defined as TMB-high (Cristescu R et al. *J Immunother Cancer* 2022, PMID: 35101941).

**Supplementary figure 2) Determination of the number of clusters**

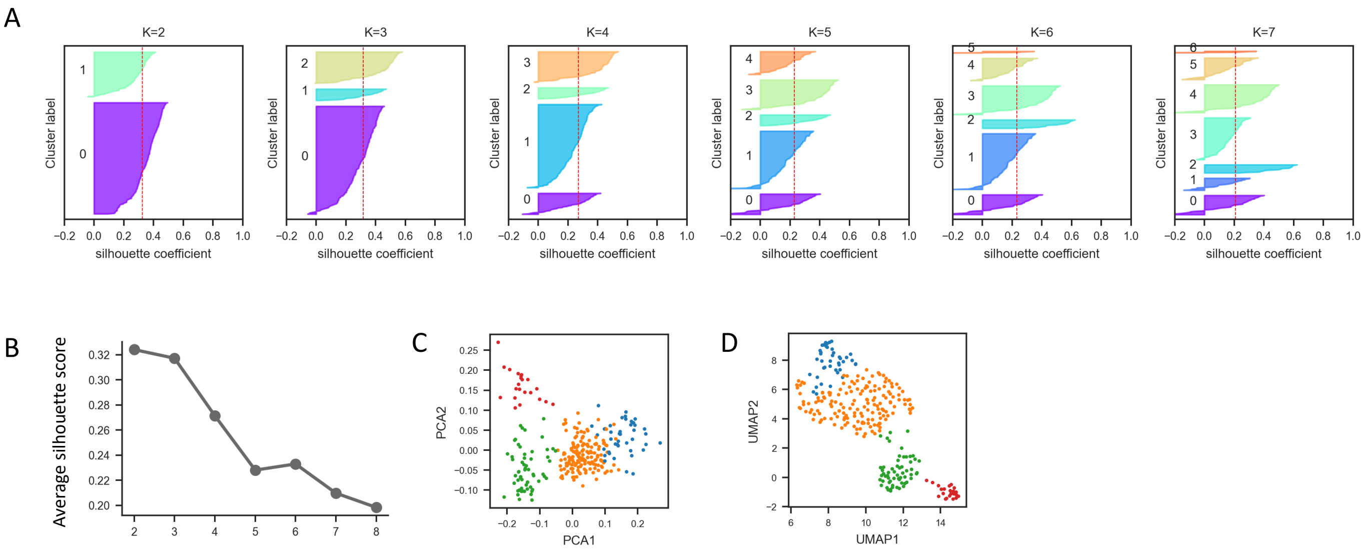


A) The silhouette plots for different numbers of clusters
The red line indicates the average silhouette score.

B) Change in the average silhouette score by number of cluster
We selected k=4 based on both the average silhouette score values and molecular differences between the assigned clusters.

C) Associations between the sample distribution and assigned subtypes using Principal Component Analysis (PCA).
The four clusters were well separated visually.

D) Associations between the sample distribution and assigned subtypes using Uniform Manifold Approximation and Projection (UMAP)
The four clusters were well separated visually.

**Supplementary figure 3) Differentially expressed gene analysis between C4 vs C1-C3 in the JGOG-TR2**

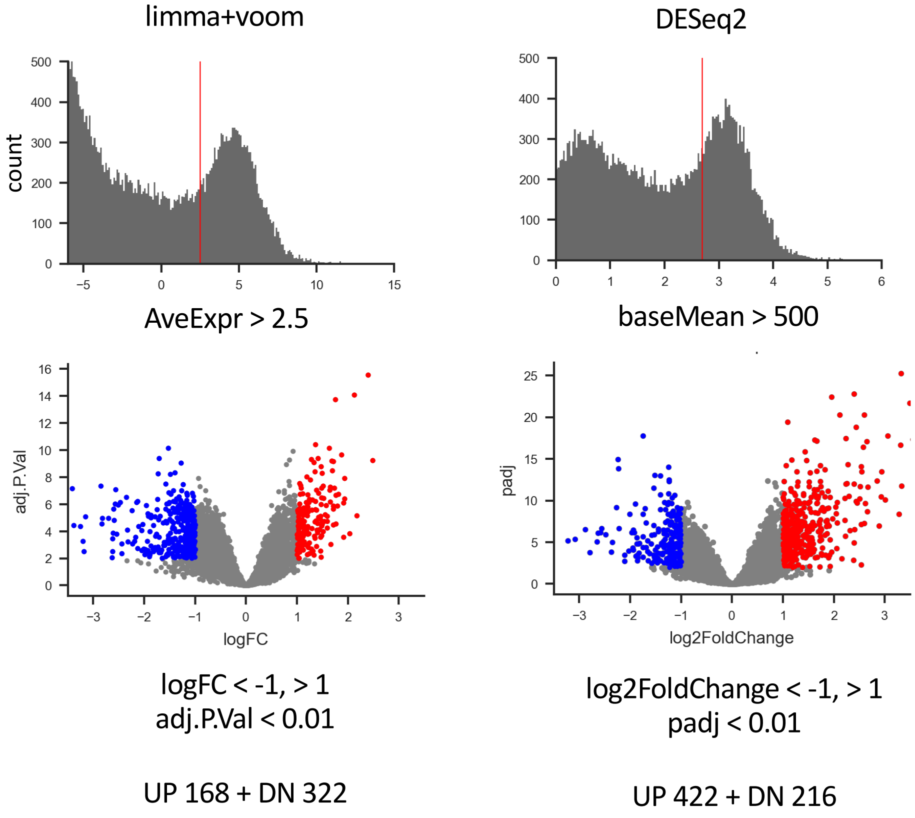


Using limma+voom (left) and DESeq2 (right), after filtering low expressed genes (AveExpr >2.5 and baseMean >500, respectively), genes with log2 fold change >1 or <-1 and False Discovery Rate < 0.01 were retained.

Upregulated 147 genes and downregulated 161 genes in C4 were selected as overlaps in the two methods and defined as JGOG-C4 gene signature UP and DN, respectively.

**Supplementary figure 4) Trend in percentage of high-grade endometrioid carcinomas among all endometrioid carcinomas of the ovary from 2000 to 2020 in the SEER database**

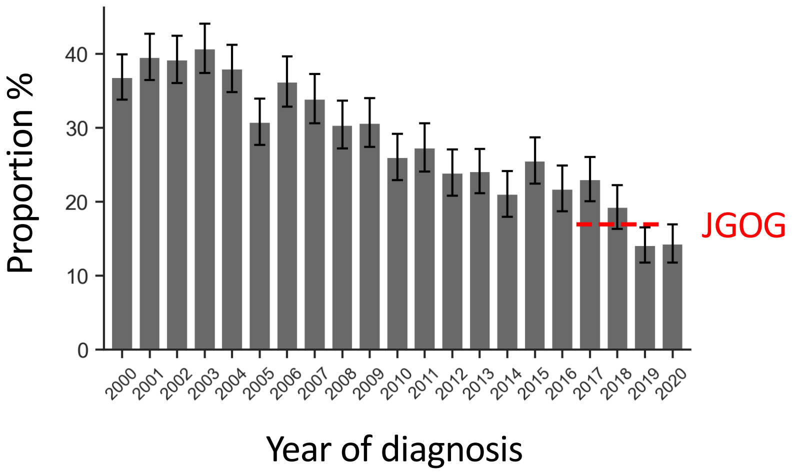


The diagnosis rate of ovarian high-grade endometroid carcinoma have decreased steadily over the two decades.

The percentage and the study period in the JGOG3025 was indicated by the red line.

Whiskers represent 95% confidence intervals.

**Supplementary figure 5) Differences in genomic profiles between HGEC and HGSC based on CPR diagnosis**

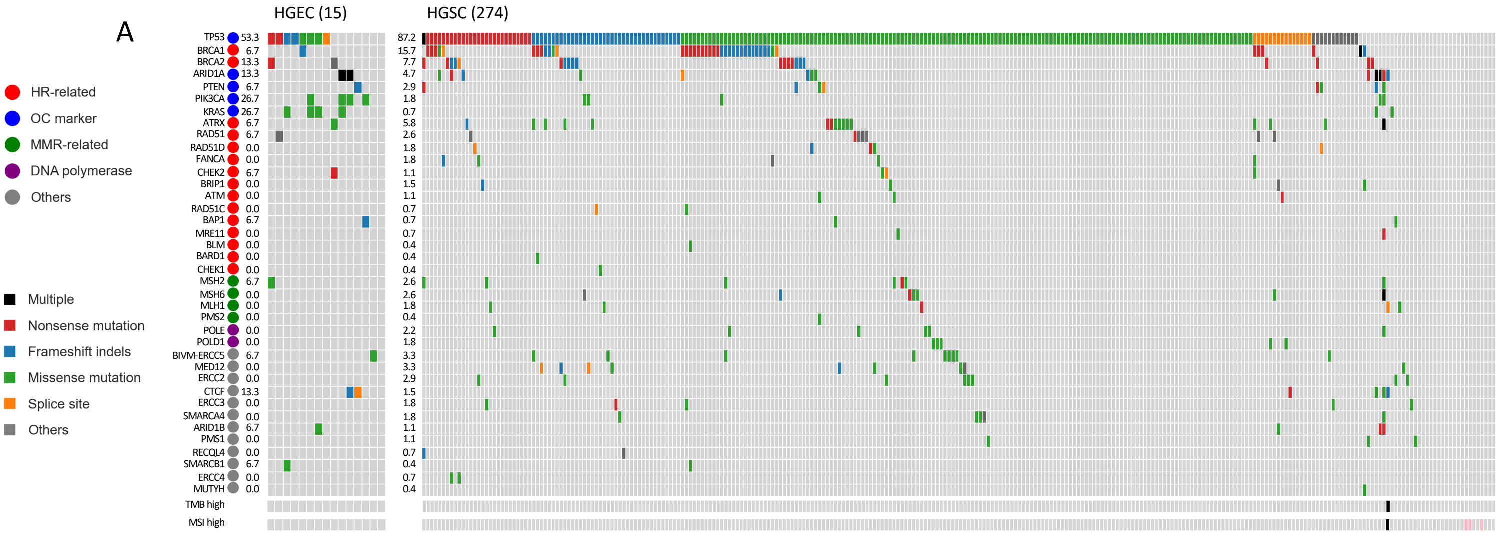


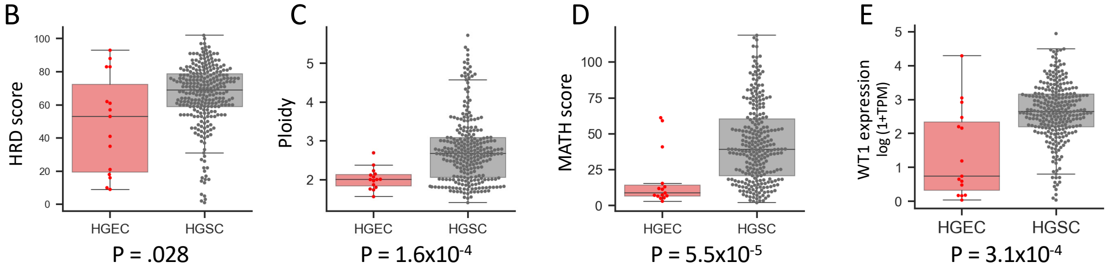


F

| | | ITI diagnosis | | |
|---|---|---|---|---|
| | | HGEC | HGSC | Others |
| CPR diagnosis | HGEC | 6 | 6 | 3 # |
| | HGSC | 10 | 258 | 6 $ |

A) Gene mutation profiles from target DNA sequencing
Mutation rates were similar to those previously reported (33020491, 34711431).

B) The HRD score was significantly higher in HGSC than HGEC

C) Estimated ploidy was significantly higher in HGSC than HGEC

D) The MATH score, an indicator of intra-tumor heterogeneity, was significantly higher in HGSC than HGEC

E) Gene expression of *WT1* was significantly higher in HGSC than HGEC

F) Discordancy in pathology diagnosis between the central pathology review (CPR) and the initial treatment institutions (ITI).
\# high-grade serous and endometroid, poorly differentiated carcinoma, undifferentiated carcinoma
$ high-grade serous and clear cell carcinoma, clear cell carcinoma (2), poorly differentiated adenocarcinoma, adenocarcinoma (non-specific) (2)

**Supplementary figure 6) Correlation between protein expression and mRNA expression of in TCGA-OV**

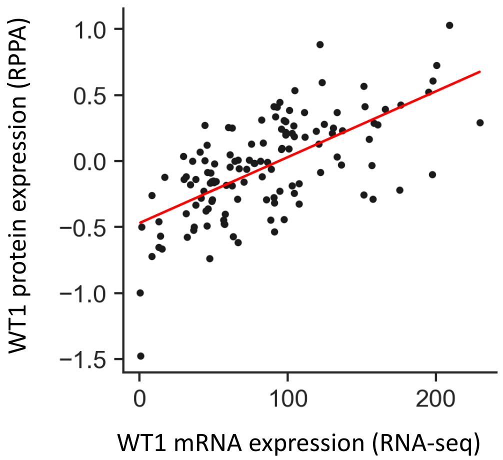


r = 0.65, P = 1.3x $10^{-16}$

*WT1* gene expression was significantly positively correlated with WT1 protein expression (Spearman correlation, r = 0.65, p = 1.3 x $10^{-16}$)

**Supplementary figure 7) Differences in molecular profiles between the four subtypes in the JGOG-TR2 cohort**

A

| | C1 | C2 | C3 | C4 | P-value | |
|---|---|---|---|---|---|---|
| Number | 41 | 160 | 59 | 22 | | |
| CPR diagnosed as HGEC | 0 | 2.5 | 8.5 | 27 | $1.8 \times 10^{-6}$ | # |
| TP53 mutation | 83 | 91 | 86 | 45 | $3.0 \times 10^{-8}$ | # |
| BRCA1/2 alteration | 2.4 | 41 | 53 | 0 | $4.0 \times 10^{-4}$ | # |
| CCNE1 amplification | 59 | 13 | 0 | 0 | .033 | # |
| ARID1A/KRAS/PIK3CA/PTEN | 7.3 | 8.1 | 5.1 | 41 | $4.2 \times 10^{-7}$ | $ |
| HRD score | 61 | 71 | 70 | 20 | $4.5 \times 10^{-45}$ | $ |
| Ploidy | 4.3 | 2.8 | 1.8 | 2.0 | $6.4 \times 10^{-5}$ | $ |
| MATH score | 49 | 44 | 37 | 24 | .0011 | $ |
| KEGG CELL CYCLE | 0.13 | 0.21 | -0.067 | -0.92 | $1.9 \times 10^{-7}$ | $ |
| G2M CHECKPOINT | 0.011 | 0.25 | -0.039 | -1.1 | $1.1 \times 10^{-9}$ | $ |
| WT1 gene expression | 2.6 | 2.7 | 2.7 | 1.5 | $8.1 \times 10^{-10}$ | $ |

% (100, 50, 0); z-score (1.5, 0.0, −1.5)

B

| | CPR HGSC | CPR HGEC | P | ITI HGSC | ITI HGEC | P | CRP/ITI HGSC | CRP/ITI HGEC | P | |
|---|---|---|---|---|---|---|---|---|---|---|
| Number | 267 | 15 | | 266 | 16 | | 257 | 25 | | |
| TP53 mutation | 87 | 53 | *** | 87 | 56 | *** | 88 | 56 | *** | # |
| BRCA1/2 alteration | 35 | 27 | | 35 | 25 | | 35 | 28 | | # |
| CCNE1 amplification | 16 | 20 | | 17 | 6.2 | | 16 | 12 | | # |
| ARID1A/KRAS/PIK3CA/PTEN | 7.9 | 47 | *** | 7.5 | 50 | *** | 6.2 | 48 | *** | # |
| HRD score | 67 | 49 | *** | 67 | 51 | ** | 67 | 51 | *** | $ |
| Ploidy | 2.8 | 2 | *** | 2.8 | 2.3 | * | 2.8 | 2.1 | *** | $ |
| MATH score | 43 | 18 | *** | 43 | 29 | | 44 | 25 | ** | $ |
| KEGG CELL CYCLE | 0.061 | -0.11 | | 0.084 | -0.47 | * | 0.085 | -0.28 | | $ |
| G2M CHECKPOINT | 0.06 | -0.1 | | 0.076 | -0.35 | | 0.079 | -0.23 | | $ |
| WT1 gene expression | 2.7 | 1.4 | *** | 2.6 | 1.9 | ** | 2.7 | 1.8 | *** | $ |

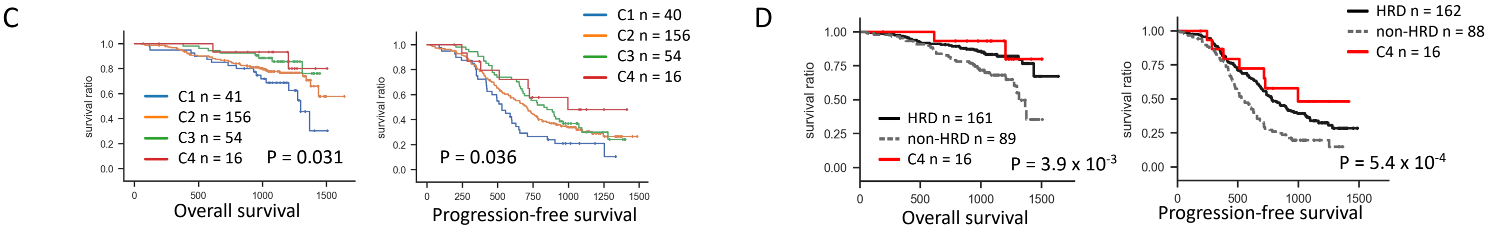


A) Summarized table of the heatmap in Figure1A.
Binary values (#) and continuous values ($) were compared between C4 and other subtypes using Chi-square test and T-test, respectively.

B) Molecular features of the tumors pathologically diagnosed as HGEC in CRP (n=15), ITI (n=16), and either of them (n=25)
Compared to the C4, significant absence of *BRCA1/2* alterations or *CCNE1* amplifications and significant difference in G2M CHECKPOINT gene expression signatures were not observed.
Statistical significance is denoted as follows: * for P < 0.05, ** for P < 0.01, and *** for P < 0.001.

C) Differences of overall (left) and progression-free (right) survival outcomes between the four subtypes in samples diagnosed as HGSC.
Even when compared within HGSC, C4 tended to have a favorable survival.

D) Differences of overall (left) and progression-free (right) survival outcomes between C4, HRD and non-HRD tumors in samples diagnosed as HGSC.
Even when compared within HGSC, C4 tended to have as a favorable survival as non-HRD.

**Supplementary figure 8) Genome-wide DNA methylation analysis associated with cell differentiation of endometrium and fallopian tubes**

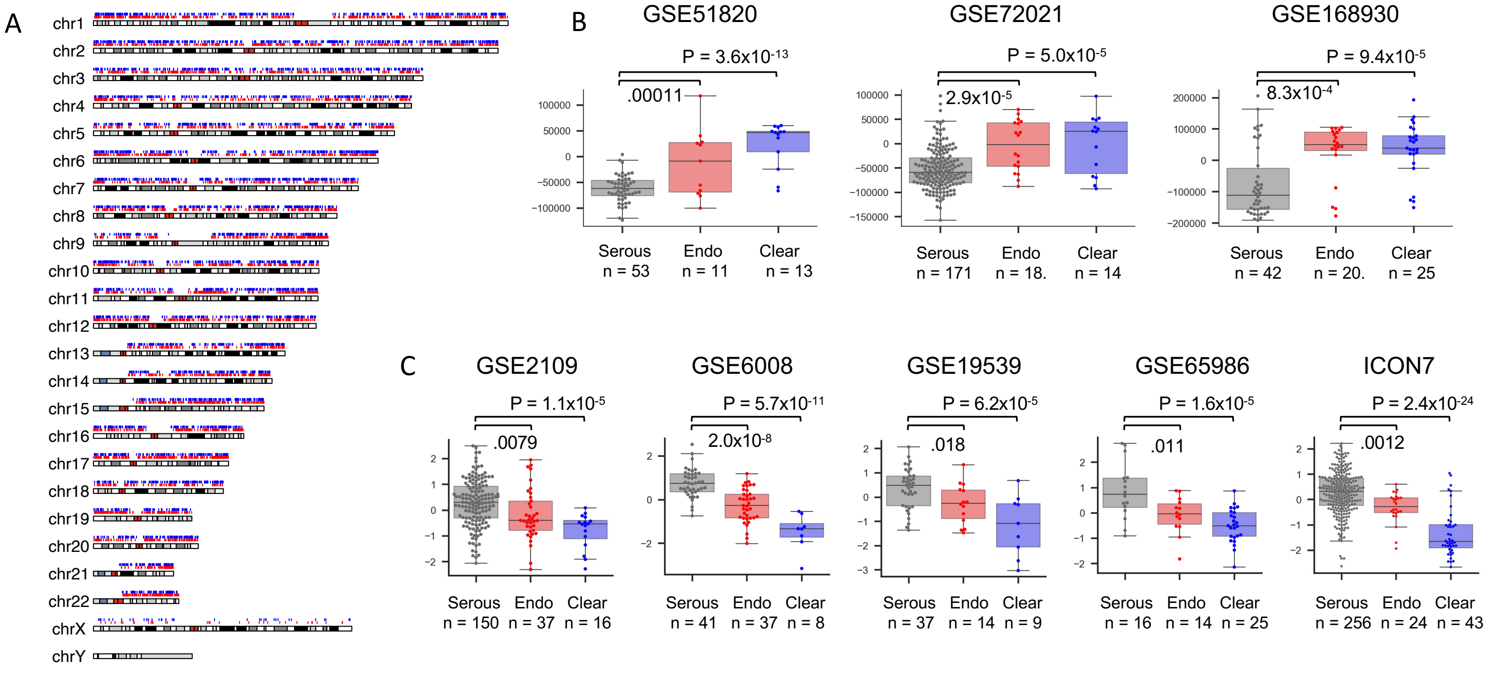


A) Differentially methylated regions between normal endometrium and fallopian tubes in GSE186458. Genomic regions that are hypermethylated in endometrium than fallopian tubes were shown in red, and the opposite in blue.

B) Comparison of EM/FT methylation scores in public ovarian cancer datasets. The scores were significantly higher in endometroid (Endo) or clear cell carcinoma (Clear) than in serous carcinoma (Serous).

C) Comparison of EM/FT silencing scores in public ovarian cancer datasets. The scores were significantly higher in Endo or Clear than in Serous.

**Supplementary figure 9) Machine learning-based tumor subtype prediction based on CNV signatures**

A

KNN

| | pC1 | pC2 | pC3 | pC4 |
|---|---|---|---|---|
| C1 | **34** | 7 | 0 | 0 |
| C2 | 0 | **160** | 0 | 0 |
| C3 | 0 | 3 | **56** | 0 |
| C4 | 0 | 0 | 1 | **21** |

Accuracy = 93.2 %

MLP

| | pC1 | pC2 | pC3 | pC4 |
|---|---|---|---|---|
| C1 | **37** | 4 | 0 | 0 |
| C2 | 1 | **159** | 0 | 0 |
| C3 | 0 | 2 | **57** | 0 |
| C4 | 0 | 0 | 2 | **20** |

Accuracy = 96.8 %

SVM

| SVM | pC1 | pC2 | pC3 | pC4 |
|---|---|---|---|---|
| C1 | **41** | 0 | 0 | 0 |
| C2 | 4 | **156** | 0 | 0 |
| C3 | 0 | 1 | **58** | 0 |
| C4 | 0 | 0 | 0 | **22** |

Accuracy = 98.2 %

XGB

| XGB | pC1 | pC2 | pC3 | pC4 |
|---|---|---|---|---|
| C1 | **37** | 4 | 0 | 0 |
| C2 | 2 | **157** | 1 | 0 |
| C3 | 0 | 6 | **53** | 0 |
| C4 | 0 | 0 | 1 | **21** |

Accuracy = 95.0 %

RFC

| RFC | pC1 | pC2 | pC3 | pC4 |
|---|---|---|---|---|
| C1 | **38** | 3 | 0 | 0 |
| C2 | 2 | **153** | 2 | 3 |
| C3 | 0 | 4 | **55** | 0 |
| C4 | 0 | 0 | 1 | **21** |

Accuracy = 96.1 %

B

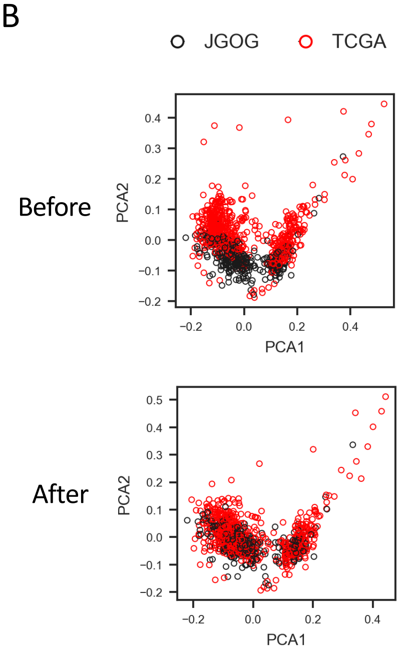


A) Internal validation using five different classifiers.
Results from leave one out validation in training data showed that all five algorithms have very high accuracy in the multi-label prediction.
KNN; K-nearest neighbor, SVM; Support vector machine, RFC; Rando`m forest, MLP; Multilayer perceptron, XGB; XGBoost
Bold letters indicate the number of matches between labels and predictions.

B) Batch correction between the JGOG-TR2 cohort (n=282) and the TCGA-OV cohort (n=555).
We used Combat_seq (33015620) for 48-channel CNV signatures to correct a batch effect.
PCA plots before (upper) and after (lower) correction showed that the distribution of the two sample groups was properly corrected.

**Supplementary figure 10) Analysis in TCGA-UCEC cohort**

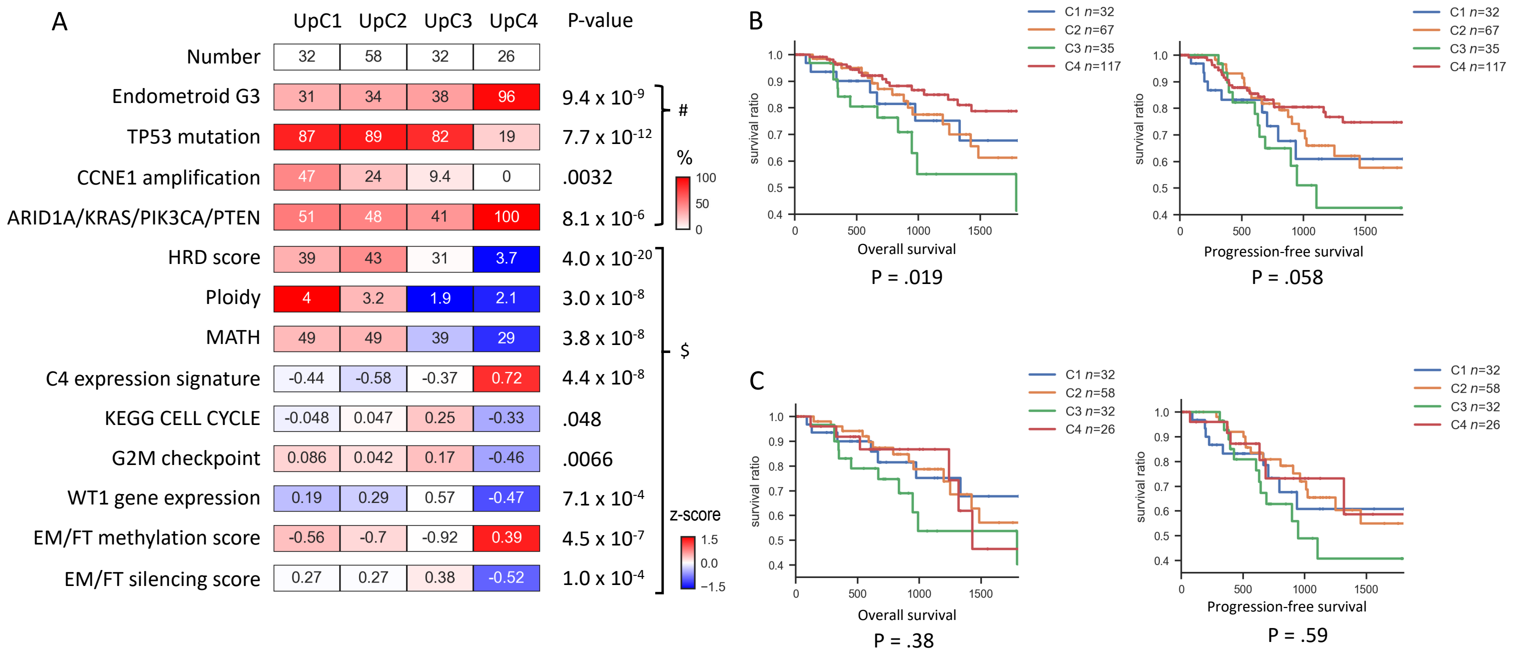


A) Differences in genomic profiles between predicted tumor subtypes after excluding POLE/dMMR tumors. Observed differences in molecular profiles among the assigned groups were similar to those in the JGOG-TR2 and TCGA-OV cohort.

B) Differences of survival outcomes between the predicted tumor subtypes
UpC4 tended to have better overall survival (left) and progression-free survival (right) than the other tumors (multivariate log rank test).

C) Differences of survival outcomes between the predicted tumor subtypes after excluding POLE/dMMR tumors
There was no significant difference among the predicted four subtypes in overall survival (left) and progression-free survival (right) (multivariate log rank test).

**Supplementary figure 11) Genome-wide DNA methylation status associated with cell differentiation of endometrium and fallopian tubes in endometrial cancer**

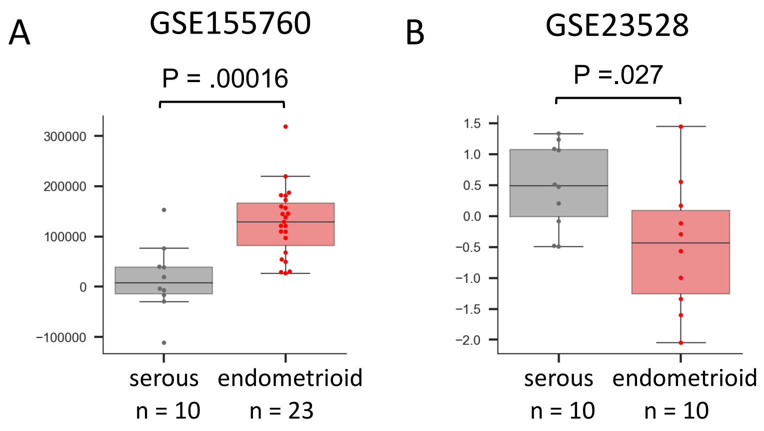


A) Comparison of the EM/FT methylation score in a public dataset of uterine endometrial cancer. In GSE155760, the scores were higher in endometrioid carcinoma than in serous carcinoma.

B) Comparison of the EM/FT silencing score in a public dataset of uterine endometrial cancer. In GSE23518, the scores were lower in endometroid carcinoma than in serous carcinoma.

**Supplementary table 1) JGOG C4 signature UP (147) and DN (161) genes**

**JGOG-C4 UP signature**

| | | |
|---|---|---|
| ABCA6 | FOSB | PLS3 |
| ABI3BP | G0S2 | PODN |
| ADAMTS4 | GADD45B | PPP1R3B |
| ADAMTS9 | GASK1B | PRICKLE2 |
| ALDH1A1 | GATM | PRKCB |
| ALDH1A3 | GLRX | PSD3 |
| ANK2 | GNAI1 | PTGIS |
| ANTXR2 | GNG2 | PTGS2 |
| AQP1 | GPC6 | PTPRE |
| ARHGAP10 | GPRASP1 | RAC2 |
| ATP2A3 | GSTM3 | RBMS3 |
| BAHCC1 | GVINP1 | RCAN1 |
| BASP1 | GYPC | RDH10 |
| BICC1 | HBB | RGS2 |
| BIRC3 | HCLS1 | RGS4 |
| BTG2 | HIC1 | RGS5 |
| C1QTNF1 | ID1 | RHOB |
| C7 | ID2 | SEMA6A |
| CACNA1H | IER3 | SERPINE2 |
| CAV1 | IGFBP4 | SERPINF1 |
| CAV2 | INPP5D | SFRP1 |
| CCDC69 | IRF8 | SLC16A7 |
| CD36 | KITLG | SLC2A3 |
| CD44 | KLF2 | SMOC2 |
| CDKN1A | KLF4 | SORBS2 |
| CELF2 | LAMA2 | STS |
| COL14A1 | LHFPL6 | SULF2 |
| CPED1 | LIF | SYNPO2 |
| CPXM2 | LMOD1 | TFPI |
| CR1 | LRRK2 | TGFA |
| CRYL1 | MAOA | TGFB3 |
| CTSH | MAOB | TINAGL1 |
| DCLK1 | MAPK10 | TLR4 |
| DLC1 | MEG3 | TNXB |
| DMD | MEG8 | TTN |
| DOCK5 | MGP | VGLL3 |
| DPP4 | MRC1 | VIM |
| DST | MYH11 | WNT2B |
| DUSP1 | NAV3 | ZFP36 |
| DUSP4 | NEDD9 | |
| DUSP5 | NFASC | |
| ECM2 | NLGN4X | |
| EFEMP1 | NLRP1 | |
| EGR3 | NR2F1 | |
| ELL2 | NR4A1 | |
| ENPP2 | NR4A3 | |
| EPB41L3 | PAPLN | |
| EPS8 | PARM1 | |
| F13A1 | PDE4B | |
| F3 | PDE5A | |
| FAT4 | PDGFRA | |
| FGF7 | PDK4 | |
| FLRT2 | PDLIM3 | |
| FOS | PGR | |

**JGOG-C4 DN signature**

| | | |
|---|---|---|
| AIF1L | HMGA2 | PUF60 |
| ALDH3B2 | HOXD3 | PYCR3 |
| ALOX12P2 | IFI27 | RAB6B |
| ANKRD33B | IFI44 | RAD51AP1 |
| ASF1B | IFI6 | RAD54B |
| ASPM | IFIT1 | RBM38 |
| ASS1 | IGF2BP2 | RECQL4 |
| ATAD2 | ISG15 | RFC4 |
| ATP6V1B1 | KCND2 | RIMS4 |
| AURKA | KCNK15 | RPL39L |
| BCAM | KIF15 | RSAD2 |
| BDH1 | KIF20A | RTP4 |
| BHLHE41 | KIF23 | S100A1 |
| BST2 | KIFC1 | SALL4 |
| BX322639.1 | KLHL14 | SAMD10 |
| C20orf204 | KLK5 | SCARA3 |
| CBS | KLK6 | SCNN1A |
| CCNE1 | KLK7 | SGO1 |
| CDC20 | KLK8 | SLC34A2 |
| CDCA5 | L1CAM | SLPI |
| CDK1 | LAMP3 | SMC4 |
| CENPF | LAPTM4B | SNCG |
| CEP55 | LINC01139 | SNHG4 |
| CERNA2 | LPAR3 | SPC24 |
| CHI3L1 | LY6E | SPON1 |
| CHODL | LYPD1 | SQLE |
| CLDN16 | MAL2 | ST6GALNAC5 |
| CLGN | MCM10 | SUSD4 |
| COL13A1 | MCM2 | TERC |
| CP | MELK | TFAP2C |
| CRABP2 | MPP7 | TMEFF1 |
| CTCFL | MROH6 | TNNI3 |
| CYP4B1 | MSLN | TNNT1 |
| DEPDC1 | MTBP | TPX2 |
| DHCR24 | MYBL2 | TRIP13 |
| DOK5 | NAT8L | TROAP |
| DSCC1 | NDC80 | TSPYL5 |
| E2F1 | NDUFB9 | TTK |
| E2F8 | NEK2 | UBE2C |
| EBP | NMU | USP18 |
| ECT2 | NPR1 | WT1 |
| ERCC6L | NR2F6 | WT1-AS |
| EXO1 | NRBP2 | XPR1 |
| FAM83D | NUF2 | XRCC2 |
| FOLR1 | NUS1P2 | ZNF257 |
| FOXM1 | PARD6B | ZNF334 |
| GALNT6 | POLQ | ZNF492 |
| GINS1 | PPP1R16A | ZNF503-AS2 |
| GMPR | PPP1R1B | ZNF572 |
| GPR27 | PRIMA1 | ZNF726 |
| GPRC5B | PRKCI | ZNF774 |
| GRB7 | PSAT1 | ZNF90 |
| H1-1 | PSRC1 | ZNF98 |
| HAGLR | PTGS1 | |

**Supplementary table 2) EM>FT (208) and FT>EM (106) methylation silencing genes**

**EM>FT methylation silencing genes**

| | | | |
|---|---|---|---|
| ABR | EPHA1-AS1 | MAFK | SEMA3F |
| ACAP3 | EPN3 | MAMSTR | SEPTIN10 |
| ACOT11 | ERC1 | MAPK10 | SEPTIN9 |
| ACTN1 | ERF | MAST4 | SH3BP1 |
| ADA2 | EVA1C | MGAT4B | SH3TC2 |
| AGAP3 | EXOC6 | MGC16275 | SHANK1 |
| AKNA | EYA4 | MICAL3 | SLC12A8 |
| AKR1C8P | FAM178B | MIR1295B | SLC22A11 |
| ALPK3 | FAM20C | MIR200A | SLC26A9 |
| ANGPTL6 | FBXW7 | MIR33B | SLC2A5 |
| ANKRD6 | FCGRT | MIR4632 | SLC34A1 |
| ANKRD65 | FGD2 | MIR4640 | SLC38A1 |
| AP3M2 | FGFR1 | MIR5587 | SLC43A2 |
| ARHGEF3 | FOXA2 | MIR575 | SLC45A4 |
| ASPSCR1 | FOXJ1 | MLC1 | SLC47A2 |
| ATP1A1 | FOXN1 | MPI | SND1 |
| BNC2 | FUT1 | MRC2 | SNRPF |
| C11orf80 | GAL3ST2 | MRPL38 | SORCS3 |
| C20orf204 | GDPD5 | MUCL3 | SOST |
| C21orf58 | GGT5 | MYO1C | SPHK1 |
| C6orf201 | GIT1 | NEU2 | SPTBN5 |
| C9orf92 | GJC2 | NMT1 | SRGAP3-AS3 |
| CABP4 | GLB1L2 | NRGN | ST6GALNAC6 |
| CALML4 | GNA15 | OXCT2 | STAB1 |
| CARD11 | GRAMD1A | PAFAH2 | STING1 |
| CCDC88B | GRAMD4 | PDXK | TACC1 |
| CCDC9B | HCAR1 | PHRF1 | TCIRG1 |
| CD6 | HMGCS1 | PIP4K2A | TDRD10 |
| CFB | HOTAIRM1 | PITX1 | TENM2 |
| CHRNB4 | HOXB9 | PKP3 | TET1 |
| CLEC18B | HSPB2 | PLEC | THUMPD2 |
| COL18A1-AS2 | HSPB2-C11orf52 | PLEKHB1 | TIAM1 |
| COX5B | IFT122 | PLEKHG3 | TLE5 |
| CROCC | IL12RB2 | PLEKHG5 | TLK2 |
| CRYAB | IL1R1 | PLSCR4 | TLR8-AS1 |
| CWH43 | IL1RL1 | PLXNB2 | TMEM63B |
| CXCL17 | KIAA0825 | POLE2 | TMIGD2 |
| CYB561 | KIFC3 | PPP2R2C | TMPRSS6 |
| CYBC1 | KRT15 | PPP5C | TNFSF13 |
| CYP2B7P | KRTAP10-3 | PRND | TNK2 |
| CYP2W1 | LCN10 | PRSS1 | TNS1 |
| DDR1 | LCN6 | PTPN7 | TP73-AS1 |
| DENND2A | LGALS2 | RADIL | TPPP3 |
| DES | LGI4 | RALB | TRABD |
| DLGAP4 | LILRA2 | RALBP1 | TRAF3 |
| DOC2B | LINC01133 | RAP1GAP | TRAK1 |
| DPEP1 | LINC02396 | RAPGEF1 | TSPAN8 |
| ELAPOR1 | LOC100129066 | RGS12 | WT1 |
| ELMOD2 | LOC284950 | RIPOR2 | ZFP36 |
| EME1 | LOC401557 | RNASE1 | ZFX |
| EML1 | LRIG3 | SCGB3A1 | ZNF330 |
| EP400P1 | LRTM2 | SELPLG | ZNF75D |

**FT>EM methylation silencing genes**

| | |
|---|---|
| ABCC1 | LOX |
| ADAMTSL5 | LRMDA |
| ADARB2 | LRRC32 |
| ADD1 | MAGI1 |
| AKNAD1 | MAK16 |
| ANK1 | MBNL2 |
| ANK2 | MIR3193 |
| ANTXR2 | MIR5188 |
| APBA2 | MMP21 |
| ARHGAP20 | MTUS1 |
| ARHGAP24 | MYT1L |
| ARL9 | NACA |
| ARRDC2 | NDUFC2 |
| ATP11A | NDUFC2-KCTD14 |
| BCR | NPLOC4 |
| BRD1 | P2RX7 |
| C1orf53 | PCLAF |
| C7orf77 | PDE7A |
| CD36 | PHETA1 |
| CDCA7 | PHF11 |
| CLCC1 | PHYHD1 |
| CLDN14 | PLXNA4 |
| CPN1 | POLR2D |
| CSGALNACT1 | QTRT1 |
| CYP2R1 | RBL2 |
| DACT1 | RGMA |
| DBR1 | RPS14P3 |
| DENND3 | SELENOW |
| DHRS7B | SKIL |
| DHRS9 | SLC35F6 |
| DIPK1A | SLC45A4 |
| DNAJC15 | SLC49A3 |
| EML6 | SLC51B |
| FAM3C | SLC6A12 |
| FOXM1 | SPATA41 |
| GAPVD1 | STC2 |
| GDPD4 | STK3 |
| GNG11 | TBRG1 |
| GRN | TEX19 |
| HAMP | TEX47 |
| HAVCR1 | TINAGL1 |
| HEYL | TMEM140 |
| HPGD | TMEM37 |
| IDNK | TUBGCP2 |
| IZUMO1R | TXNDC11 |
| KIF25 | UBC |
| LINC00161 | WNK2 |
| LINC00310 | XBP1 |
| LINC00460 | ZBTB16 |
| LOC100287010 | ZDHHC11 |
| LOC100289561 | ZNF14 |
| LOC100506422 | ZNF765 |
| LOC100630923 | ZNF821 |

**Supplementary table 3) List of publicly available datasets**

| Datasets | Name, ID | PMID | URL |
|---|---|---|---|
| Whole genome bisulfate sequencing | GSE186458 | 36599988 | https://www.ncbi.nlm.nih.gov/geo/query/acc.cgi?acc=GSE186458 |
| DNA methylation array analysis | GSE51820 | 24382740 | https://www.ncbi.nlm.nih.gov/geo/query/acc.cgi?acc=GSE51820 |
| | GSE72021 | 26629914 | https://www.ncbi.nlm.nih.gov/geo/query/acc.cgi?acc=GSE72021 |
| | GSE155760 | 32817081 | https://www.ncbi.nlm.nih.gov/geo/query/acc.cgi?acc=GSE155760 |
| | GSE168930 | 34294135 | https://www.ncbi.nlm.nih.gov/geo/query/acc.cgi?acc=GSE168930 |
| | GSE226823 | 37874327 | https://www.ncbi.nlm.nih.gov/geo/query/acc.cgi?acc=GSE226823 |
| Gene expression analysis | GSE2109 | - | https://www.ncbi.nlm.nih.gov/geo/query/acc.cgi?acc=gse2109 |
| | GSE6008 | 16452189 | https://www.ncbi.nlm.nih.gov/geo/query/acc.cgi?acc=GSE6008 |
| | GSE19539 | 20386695 | https://www.ncbi.nlm.nih.gov/geo/query/acc.cgi?acc=GSE19539 |
| | GSE44104 | 23934190 | https://www.ncbi.nlm.nih.gov/geo/query/acc.cgi?acc=GSE44104 |
| | GSE65986 | 26147301 | https://www.ncbi.nlm.nih.gov/geo/query/acc.cgi?acc=GSE65986 |
| | ICON7 | 33149148 | https://ega-archive.org/studies/EGAS00001003487 |
| | GSE23518 | 21079744 | https://www.ncbi.nlm.nih.gov/geo/query/acc.cgi?acc=GSE23518 |
| TCGA multi-omics<br>• Somatic gene mutation<br>• SNP array-based CNV<br>• RNA-seq gene expression<br>• DNA methylation array | TCGA-OV<br>TCGA-UCEC | 21720365<br>23636398 | GDC Data Portal v1 (https://portal.gdc.cancer.gov/v1) :Data Type<br>• Masked Somatic Mutation<br>• Allele-specific Copy Number Segment<br>• Gene Expression Quantification<br>• Methylation Beta Value |
| • Pathogenic germline mutation (controlled access) | phs000178 (dbGaP) | 29625052 | https://gdc.cancer.gov/about-data/publications/PanCanAtlas-Germline-AWG |
